# Genome-Wide Discovery Reveals Adipose-Specific and Systemic Regulators of Insulin Resistance

**DOI:** 10.64898/2026.03.31.26349822

**Authors:** Mario Garcia-Urena, Pearlyn Jia Ying Toh, Raquel Sanz Martinez, Rama Kaalia, Mayank Murali, Hesam Dashti, Yaxin Jing, César Cunha, María Jose Romero-Lado, Yuejiao Huang, Martin Wabitsch, Melina Claussnitzer, Tuomas O. Kilpeläinen

## Abstract

Insulin resistance (IR) is a key driver of cardiometabolic disease, yet its genetic and regulatory architecture remains incompletely understood. We performed a multi-trait GWAS of fasting insulin, triglycerides, and HDL cholesterol (n ≤ 1.25 million), identifying 282 IR-associated loci, including 70 novel. Polygenic score analyses linked IR to an adverse fat distribution characterized by reduced subcutaneous and increased visceral and ectopic fat. Stratifying loci by BMI associations revealed biologically distinct variant subgroups with divergent regulatory activity during adipogenesis. Enhancer-to-gene mapping implicated 72 loci in adipose-specific regulation, including a novel *LAMB1* locus, wherein knockdown enhanced adipogenesis in vitro. Coding variants in *PLAUR* and *INPP5A* implicated inflammatory and calcium signaling pathways, while Mendelian Randomization identified circulating KLK1 as a candidate causal mediator in hyperinsulinemia. Our findings refine the genetic landscape of IR, highlight adipose dysfunction as a central mechanism, and nominate new targets for mechanistic and therapeutic investigation.

---

Insulin resistance (IR), a hallmark of metabolic dysfunction, arises from impaired insulin signaling in key metabolic tissues, particularly adipose tissue, liver, and skeletal muscle**^1^**. Although IR is a central pathogenic mechanism underlying type 2 diabetes, cardiovascular disease, and metabolic dysfunction-associated steatotic liver disease, no currently approved pharmacological treatments directly restore insulin sensitivity in insulin-responsive tissues**^1^**. Existing treatments, such as metformin and GLP-1 receptor agonists, primarily improve glycemia and promote weight loss, but exert only moderate effects on insulin sensitivity, leaving the fundamental defect in insulin action unresolved. This therapeutic gap underscores the need for novel strategies grounded in a deeper understanding of the molecular mechanisms that regulate insulin responsiveness.

While IR is commonly associated with obesity and weight gain, it is now increasingly recognized as a consequence of adipose tissue dysfunction rather than of excess adiposity alone^2,3^. This dysfunction is linked to pathological remodeling of adipose tissue, including adipocyte hypertrophy^4^, impaired adipogenesis^5^, immune cell infiltration^6^, and fibrosis^7^, which collectively disrupt systemic metabolic homeostasis and contribute to IR^8^. Yet, the causal molecular mechanisms that predispose individuals to adipose tissue dysfunction, and thereby to IR, remain incompletely understood.

Human genetic variants provide a powerful lens to dissect the biology of IR, serving as natural perturbations that reveal causal pathways in insulin action. Genome-wide association studies (GWAS) of IR-related traits, such as fasting insulin (FI) and homeostasis model assessment of IR (HOMA-IR), have identified loci enriched for regulatory activity in adipose tissue^9,10^. However, these studies have been constrained by modest sample sizes, restricting deeper insights into tissue-specific mechanisms. Multi-trait GWAS approaches have begun to overcome this limitation. A landmark study by Lotta et al. combined BMI-adjusted FI (FI_adjBMI_), triglycerides (TG), and high-density lipoprotein (HDL) cholesterol to identify 53 IR loci, many exhibiting lipodystrophy-like features characterized by reduced total body fat and dysfunctional adipose tissue^11^. Subsequent work by Oliveri et al. and DeForest et al. showed that TG/HDL ratio serves as a robust proxy for IR, particularly within adipose tissue, capturing related metabolic perturbations such as impaired de novo lipogenesis and increased lipolysis^12,13^. Nonetheless, the genetic and regulatory architecture of IR and its relationship to adipose tissue dysfunction remain incompletely defined.

In this study, we perform a novel multi-trait GWAS integrating the latest and largest GWAS for FI_adjBMI_, TG, and HDL cholesterol, identifying 282 independent loci associated with IR, including 70 previously unreported signals. Polygenic risk score (PRS) analyses show that the loci collectively contribute to an adverse fat distribution characterized by reduced subcutaneous and increased visceral and ectopic fat. Classification of loci according to their BMI associations further reveals variant subgroups with distinct effects on fat distribution and ectopic fat accumulation, as well as divergent gene regulatory signatures during adipogenesis. Through enhancer-promoter mapping, we prioritize adipose-specific candidate genes and enhancers for 72 loci, uncovering regulatory mechanisms that may play direct roles in adipose tissue dysfunction and IR. Moreover, we identify coding variants and circulating proteins with potential causal contributions to IR. Collectively, our findings provide a comprehensive view of the genetic landscape underlying IR and uncover both adipose-specific and systemic regulatory pathways that may inform future therapeutic strategies.

## Results

### Study overview

To comprehensively define the genetic and regulatory architecture of insulin resistance (IR), we implemented a multi-step analytical framework (**Fig. 1**). First, we performed multi-trait association analyses by leveraging GWAS of European ancestry for clinical surrogates of IR: FI_adjBMI_ (n_max_=151,031)^10^, HDL cholesterol (n_max_=1,244,580)^14^, and TG (n_max_=1,253,277)^14^. We then characterized the aggregated effects of identified loci through PRS analyses across anthropometric and cardiometabolic traits. To assess whether IR loci were enriched for regulatory programs active in specific tissues and cell types, we performed gene and regulatory enrichment. We further refined these analyses by stratifying loci according to their BMI associations to identify distinct variant subgroups.

**Fig. 1:**
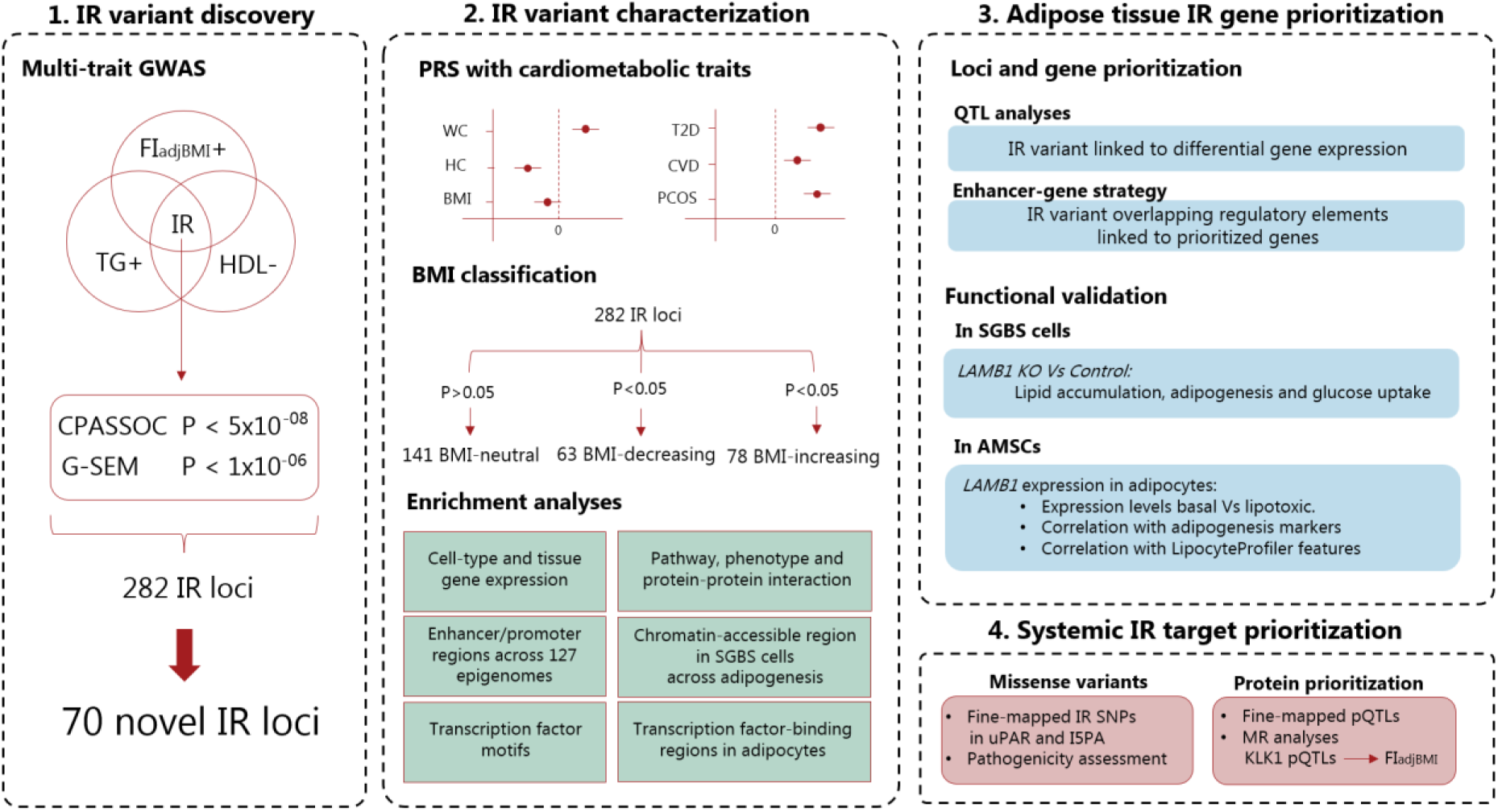
Overview of study design. **(1)** IR variant discovery: IR-associated variants were identified by integrating FI_adjBMI_, HDL cholesterol, and TG using two complementary multi-trait GWAS approaches: CPASSOC^15^ and G-SEM^16^. **(2)** IR variant characterization: PRS and enrichment analyses were performed across all loci and three BMI-defined subgroups (141 BMI-neutral, 63 BMI-decreasing, 78 BMI-increasing). **(3)** Adipose tissue IR gene prioritization: *cis*-eQTL, *cis*-sQTL, and enhancer-to-gene mapping in mature adipocytes were used to identify loci with adipose candidate genes. Further *in vitro* functional follow-up was performed on *LAMB1*. **(4)** Systemic IR target prioritization: Fine-mapped coding variants were assessed for pathogenicity and *cis*-pQTL-linked circulating proteins were evaluated for causal effects on FI_adjBMI_. **AMSC**: adipose-derived mesenchymal stem cells; **BMI**: body mass index; **CPASSOC**: Cross-Phenotype Association Analysis Using Summary Statistics; ***cis*-eQTL**: *cis-*expression quantitative trait locus; ***cis*-pQTL**: *cis-*protein quantitative trait locus; ***cis*-sQTL**: *cis-*splicing quantitative trait locus; **FI_adjBMI_**: BMI-adjusted fasting insulin: **G-SEM:** Genomic Structural Equation Modeling; **HDL**: high-density lipoprotein; **PRS:** polygenic risk scores; **SGBS:** Simpson-Golabi-Behmel syndrome; **TG**: triglycerides.

To prioritize adipose-relevant IR genes, we integrated *cis*-expression quantitative trait loci (*cis*-eQTL) and *cis*-splicing QTL (*cis*-sQTL) data from subcutaneous (SAT) and visceral adipose tissue (VAT) and performed enhancer-to-gene mapping in mature adipocytes. We performed further functional follow-up on the novel IR target *LAMB1*, including siRNA-mediated knockdown assays and analyses of image-derived phenotypes from LipocyteProfiler screens during adipogenesis.

Finally, to identify systemic mediators of IR, we identified fine-mapped coding variants within IR loci as well as examined *cis*-protein QTL (*cis-*pQTL)-linked circulating proteins for potential causal effects on FI_adjBMI_.

### Integrative genomic analyses identify 282 genetic loci associated with insulin resistance

To identify genetic loci associated with IR, we performed a multi-trait GWAS for FI_adjBMI_, HDL cholesterol, and TG (**Supplementary Table 1**)^10,14^. Recognizing that different multi-trait GWAS methodologies have distinct statistical assumptions and potential biases, we applied two complementary approaches – Cross-Phenotype Association Analysis Using Summary Statistics (CPASSOC)^15^ and Genomic Structural Equation Modeling (G-SEM)^16^ – to ensure robustness of our association signals (**Supplementary Information, Supplementary Fig. 1**). CPASSOC (S_het_ test) uses a meta-analytic framework to detect pleiotropic effects while accounting for estimated trait correlations. Although highly sensitive to shared genetic signals, it does not model a latent genetic factor underlying the traits. In contrast, G-SEM explicitly estimates a latent genetic component by modeling the genetic covariance structure across traits, enabling GWAS of this shared factor. To minimize false positives, we required loci to reach P<5x10^-8^ in CPASSOC and replicate at P<1x10^-6^ in G-SEM (**Methods, Supplementary Information**). Furthermore, we only included variants with consistent IR effect directions (i.e. FI_adjBMI_-increasing, TG-increasing, HDL-decreasing) and excluded those showing significant heterogeneity across traits (G-SEM Q heterogeneity test P<5x10^-8^) (**Supplementary Table 2**).

We identified 282 IR-associated loci, 70 of which have not been previously reported in association with IR traits^9,10-13,17-20^ and are thus considered novel (**Supplementary Table 2**). Colocalization analysis using HyPrColoc^21^ revealed that 57 of the 70 novel IR loci colocalized (posterior probability >0.30) with sub-genome-wide significant (P>5x10^-8^) signals for TG/HDL ratio^12^, suggesting that our multi-trait GWAS framework incorporating FI_adjBMI_ enhanced power to detect these associations (**Supplementary Table 2**). Importantly, 62 of the 282 IR loci (6 of the 70 novel loci) also reached Bonferroni-corrected significance (P_CPASSOC_ < 0.05/282) in analyses of non-European ancestries (African, East Asian, Hispanic and/or South Asian) ^10, 14^, despite smaller sample sizes (n = 8,101 to 119,298) (**Supplementary Table 2**), supporting the cross-population generalizability of these findings.

### Polygenic score and enrichment analyses highlight adipose tissue’s central role in IR

To assess the cumulative impact of the 282 IR loci on complex traits and disease risk, we constructed a summary statistics-based polygenic risk score (PRS)^22^, weighting alleles by effect sizes derived from the G-SEM common factor GWAS. As expected, the PRS was robustly associated with the three primary fasting IR traits used in our multi-trait GWAS, as well as with other established IR traits, including TG/HDL ratio^12^ and Stumvoll insulin sensitivity index (ISI_adjBMI_)^23^ (**Fig. 2A**).

**Fig. 2:**
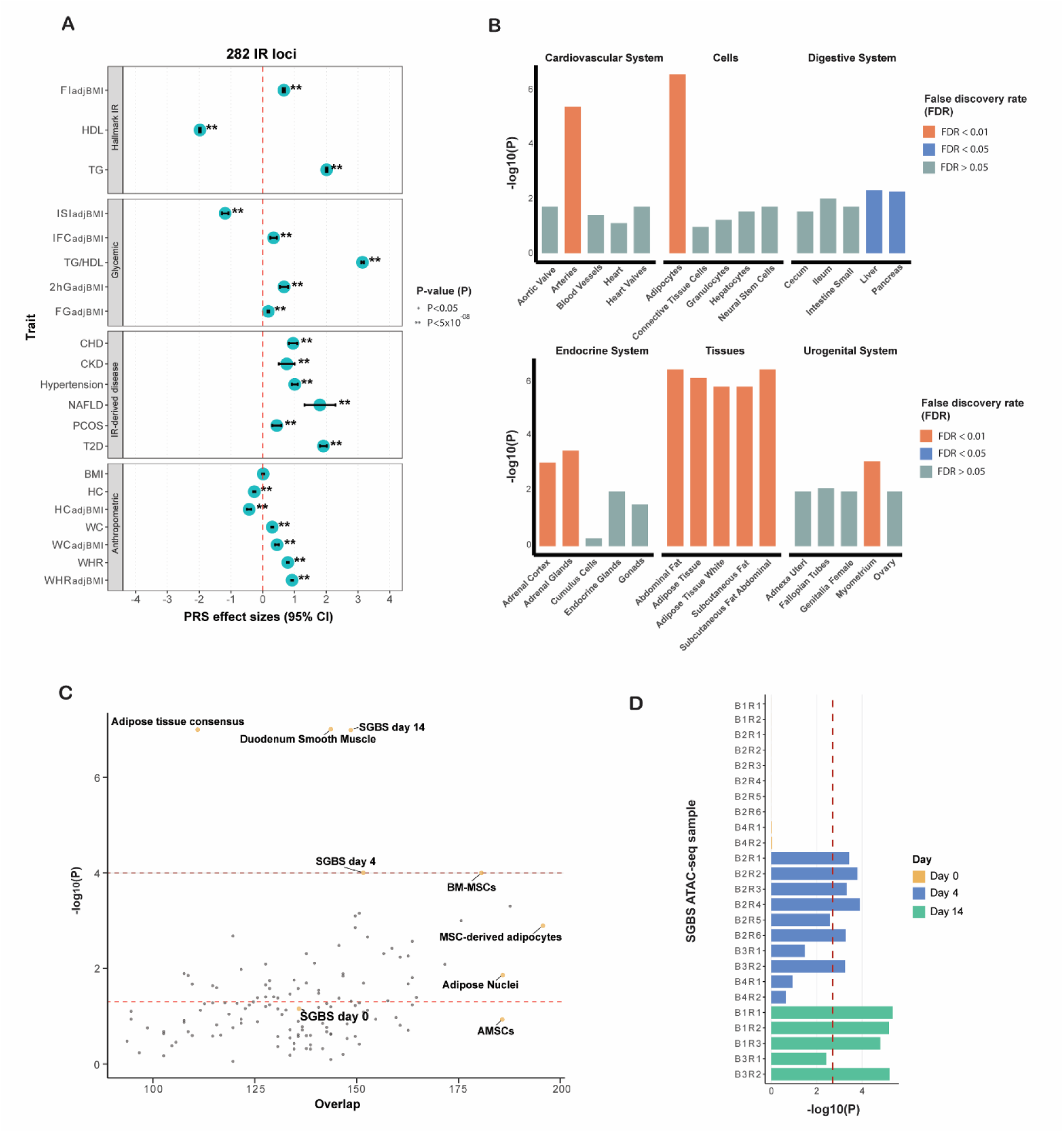
**A)** Associations of 282 IR loci with glycemic traits, anthropometric traits, and IR-related diseases. **B)** DEPICT tissue enrichment results. FDR < 0.01 in red, FDR < 0.05 in blue, and FDR > 0.05 in grey. **C)** GoShifter enrichment results for active enhancer and promoter regions from 15-state chromatin data for all tissues and cell types from ROADMAP, chromatin accessibility consensus peaks from METSIM bulk adipose tissue biopsies, and chromatin accessibility consensus peaks from SGBS cells at days 0, 4 and 14. **D)** CHEERS enrichment for consensus SGBS chromatin accessibility peaks at day 0 (orange), day 4 (blue), and day 14 (green) across ATAC-seq samples. Samples are labelled by batch and replicate (e.g., B1R1 = batch 1, replicate 1). **AMSCs:** adipose-derived mesenchymal stem cells; **BM-MSCs:** bone marrow-derived mesenchymal stem cells; **CI**: confidence intervals; **FI_adjBMI_**: BMI-adjusted fasting insulin; **HDL**: High density lipoprotein cholesterol; **TG**: triglycerides; **ISI_adjBMI_**: BMI-adjusted Stumvoll insulin sensitivity index; **TG/HDL**: TG-HDL ratio; **2hG_adjBMI_**: BMI-adjusted two-hour glucose; **FG_adjBMI_**: BMI-adjusted fasting glucose; **CHD**: coronary heart disease; **CKD**: chronic kidney disease; **NAFLD**: non-alcoholic fat liver disease; **PCOS**: polycystic ovarian syndrome; **SGBS**: Simpson-Golabi-Behmel syndrome; **T2D**: type 2 diabetes; **BMI**: body mass index; HC: hip circumference; **HC_adjBMI_**: BMI-adjusted hip circumference; **WC**: waist circumference; **WC_adjBMI_**: BMI-adjusted waist circumference; WHR: waist-hip ratio; **WHR_adjBMI_**: BMI-adjusted waist-hip ratio.

We next assessed PRS associations across anthropometric traits, imaging-derived fat distribution measures and IR-related disease outcomes (**Supplementary Table 1**)^24-30^. The IR PRS was associated with an unfavorable fat distribution phenotype, characterized by increased waist circumference and visceral adipose tissue (VAT) volume, and ectopic fat accumulation in liver and muscle, alongside decreased hip circumference and gluteofemoral subcutaneous adipose tissue (GSAT) volume (**Fig. 2A, Supplementary Table 3**). Notably, these associations were not accompanied by an increase in BMI (**Fig. 2A**). The PRS was also associated with an increased risk for multiple IR-related diseases, including T2D, coronary heart disease (CHD), chronic kidney disease (CKD), hypertension, non-alcoholic fat liver disease (NAFLD), and polycystic ovarian syndrome (PCOS) (**Fig. 2A, Supplementary Table 3**). These findings were consistent across both weighted and unweighted PRS models (**Supplementary Table 3**).

To identify tissues enriched for regulatory activity at IR-associated loci, we performed Data-driven Expression Prioritized Integration for Complex Traits (DEPICT) enrichment analysis^31^. The strongest enrichment signals were observed in white adipose tissue, abdominal fat, abdominal subcutaneous fat, and adipocytes (FDR < 0.01). Additional significant enrichments were found in arteries, adrenal gland, serous membrane of the adrenal cortex, and uterine myometrium. At FDR<0.05, liver and pancreas also showed significant enrichment (**Fig. 2B; Supplementary Table 4**). Gene set enrichment analysis implicated 771 significantly enriched pathways, phenotypes, and protein interactions at FDR<0.05 (**Supplementary Table 5**), which clustered into three major biological modules: hyperglycemia and dyslipidemia, cell membrane interactions, and transcriptional regulation (**Supplementary Fig. 5, Supplementary Table 6**).

To further characterize regulatory architecture, we applied GoShifter^32^ on ROADMAP’s 15-core chromatin state annotations across 127 epigenomes^33^. Although significant enrichment (P<1x10^-4^) was detected only in promoter and enhancer regions of duodenum smooth muscle, adipose-related epigenomes presented the highest overlap with IR loci (**Fig. 2C, Supplementary Table 7**). Analysis of METSIM’s consensus ATAC-seq peaks from 11 bulk abdominal subcutaneous adipose tissue (ASAT) samples**^34^** confirmed significant enrichment in active enhancers and promoters of adipose tissue (P<1x10^-4^).

To dissect regulatory dynamics during adipogenesis, we analyzed ATAC-seq data from Simpson-Golabi-Behmel syndrome (SGBS)^35^ cells at three differentiation timepoints: day 0 (preadipocytes), day 4 (immature adipocytes), and day 14 (mature adipocytes)^34^. GoShifter revealed strongest enrichment in mature adipocytes (P<1x10^-4^), moderate enrichment in immature adipocytes (P=1x10^-4^), and no enrichment in preadipocytes (P=0.07) (**Fig. 2C**). Chromatin Element Enrichment Ranking by Specificity (CHEERS) enrichment analysis^36^ corroborated these findings, showing significant enrichment (P<0.05/25) in consensus accessible regions in mature and immature adipocytes, but not in preadipocytes (**Fig. 2D, Supplementary Table 8**). Transcription factor (TF) binding enrichment analyses in adipocytes and SGBS cells, computed using chromatin immune-precipitation atlas (ChIPatlas)**^37^**, further highlighted differentiation stage-specific regulators, including CEBPA, CEBPB, E2F4, PPARG, RELA, NR3C1, BRD4, and MED1, which were enriched at accessible IR loci in immature and mature adipocytes (FDR<0.05), but not in preadipocytes (**Supplementary Table 9**). Taken together, these findings suggest that IR loci exert stage-specific regulatory effects during adipogenesis, engaging regulatory programs after adipocyte commitment rather than in undifferentiated preadipocytes.

### BMI-stratified analysis of IR loci reveals distinct adipose biology and regulatory programs

Although IR is commonly linked to obesity^3^, the PRS derived from the 282 IR loci showed no significant association with BMI (**Fig. 2A**). This finding is inconsistent with prior reports indicating that many IR loci exhibit lipodystrophy-like effects, often characterized by reduced BMI^11^. We hypothesized that stratifying IR loci based on their direction of effect on BMI (n_max_= 806,834)^24^ could reveal biologically distinct mechanisms contributing to adipose dysfunction.

We classified the 282 IR loci into three subgroups: 141 BMI-neutral (P>0.05), 63 BMI-decreasing (P<0.05), and 78 BMI-increasing (P<0.05) loci. Despite their divergent effects on BMI, PRS analyses showed that all three subgroups were consistently associated with an adverse fat distribution phenotype, characterized by decreased GSAT volume but increased VAT volume and liver fat (**Supplementary Table 3**). These shared associations underscore subcutaneous adipose tissue dysfunction and visceral and ectopic fat accumulation as core mechanisms underlying IR. All three subgroups were also significantly associated with increased risk of IR-related diseases: T2D, CHD, CKD, hypertension, NAFLD, and PCOS (**Supplementary Table 3**).

To investigate tissue-level regulatory differences between subgroups, we performed enrichment analyses using DEPICT. BMI-neutral and BMI-decreasing IR loci were significantly enriched for regulatory activity in adipose tissue (FDR<5%), whereas BMI-increasing loci showed no significant enrichment (**Supplementary Table 4**). This suggests that BMI-neutral and BMI-decreasing loci may contribute to IR by acting directly on adipose tissue function, whereas BMI-increasing loci may act via other tissues.

Focusing on the adipose-enriched BMI-neutral and BMI-decreasing subgroups, we observed that both groups of loci were associated with reduced GSAT and increased VAT volume, yet only the BMI-decreasing loci were additionally associated with reduced ASAT volume and increased muscle fat infiltration (**Fig. 3A**). This pattern implicates BMI-decreasing IR loci to a broader subcutaneous adipose dysfunction and lipid spillover phenotype.

**Fig. 3:**
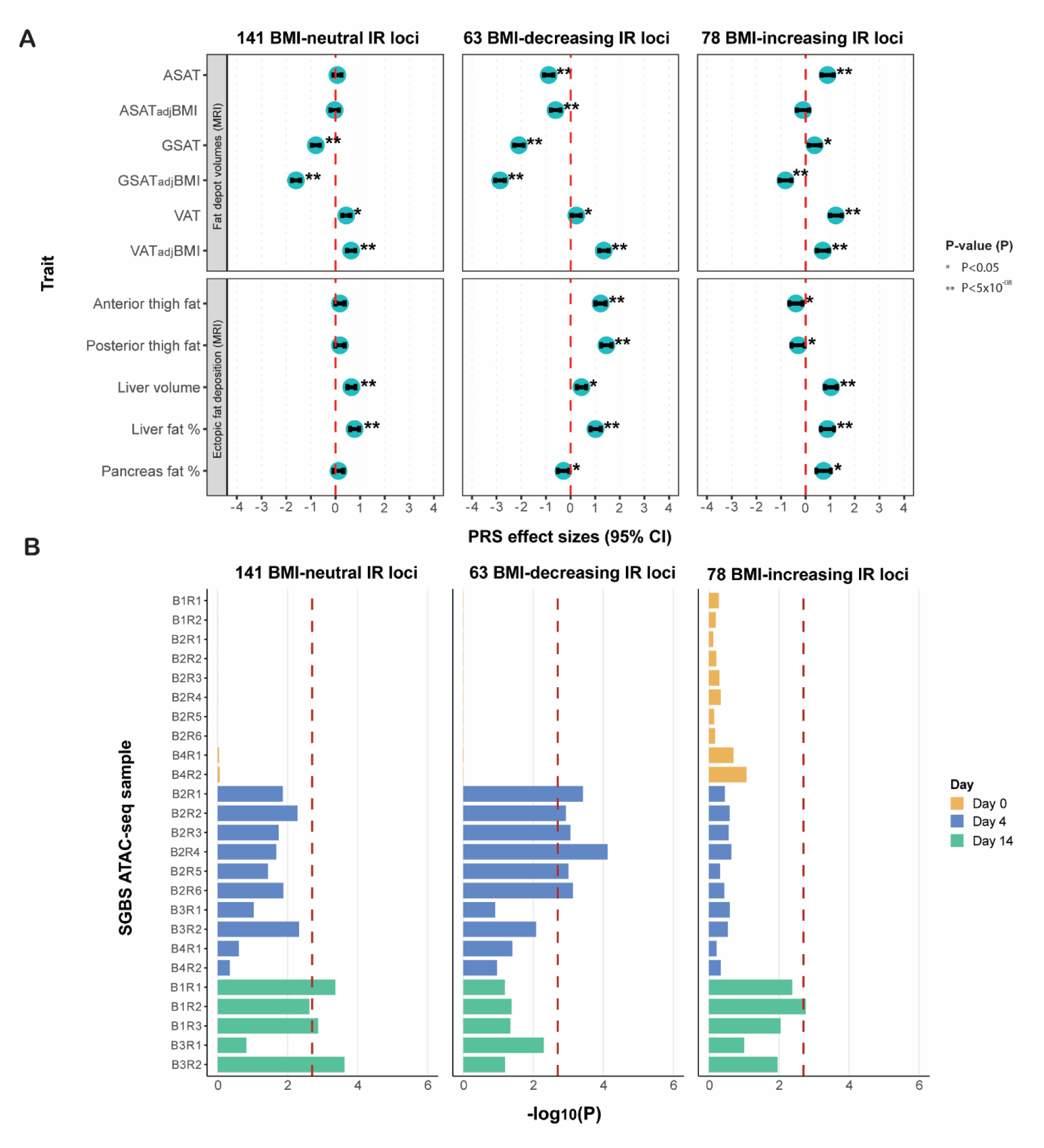
Characterization of BMI-neutral, BMI-decreasing and BMI-increasing IR loci. **A)** Associations with MRI-derived adipose tissue volume and ectopic fat accumulation traits. **B)** CHEERS enrichment results for chromatin accessibility peaks at day 0 (orange), day 4 (blue) and day 14 (green) across SGBS ATAC-seq samples from Perrin et al^28^. Samples are labelled by batch and replicate (e.g., B1R1 = batch 1, replicate 1). **ASAT**: abdominal subcutaneous adipose tissue; **ASAT_adjBMI_**: BMI and height adjusted ASAT; GSAT: gluteofemoral subcutaneous adipose tissue; **GSAT_adjBMI_**: BMI and height adjusted GSAT, **PRS**: polygenic risk scores; **SGBS**: Simpson-Golabi-Behmel syndrome; **VAT**: visceral adipose tissue; **VAT_adjBMI_**: BMI and height adjusted VAT.

Regulatory enrichment analyses using ROADMAP chromatin-state data across 127 epigenomes and ATAC-seq peaks from 11 ASAT samples in METSIM showed nominal adipose tissue enrichment (P<0.05) for both BMI-neutral and BMI-decreasing loci (**Supplementary Table 7**). CHEERS results revealed that BMI-decreasing loci were enriched in accessible chromatin regions at day 4 of SGBS cell differentiation (P<0.05/25), whereas BMI-neutral loci were enriched at day 14 (**Fig. 3B, Supplementary Table 8**), suggesting that the former may act during early adipogenesis, whereas the latter exert effects in mature adipocytes.

TF binding enrichment analyses using ChIPAtlas further supported this model. At day 4, the 63 BMI-decreasing lead variants or proxies (r^2^>0.8) were significantly enriched for CEBPB and MED1 binding (FDR<0.05), key regulators of early adipogenesis (**Supplementary Table 9**). At day 14, BMI-neutral loci were enriched for MED1, CEBPA, PPARG, RELA, and BRD4, involved in transition to mature adipocytes (**Supplementary Table 9**)^38^. TF motif enrichment analyses using Hypergeometric Optimization of Motif EnRichment (HOMER)^39^ also showed enrichment for CEBPB motifs in BMI-decreasing loci at days 4 and 14 (FDR<0.05) (**Supplementary Table 10**).

To contextualize our findings, we compared our IR subgroups with eight T2D variant clusters defined by Suzuki et al. using CHEERS.^40^ T2D loci with lipodystrophy-like effects were enriched at day 4 of adipogenesis (P<0.05/25), mirroring the pattern observed for the 63 BMI-decreasing IR loci (**Supplementary Table 8**). Conversely, T2D loci linked to metabolic syndrome-like phenotypic signatures were enriched at day 14, paralleling the BMI-neutral loci. These similarities suggest cell state-specific regulatory mechanisms between distinct IR and T2D subgroups, providing a framework for future mechanistic studies in relevant adipose contexts.

### Enhancer-promoter interactions link IR loci to adipocyte gene regulation

To elucidate the regulatory mechanisms by which IR loci influence adipose tissue biology, we first assessed *cis*-regulatory effects on gene expression and alternative splicing in ASAT by examining conditionally independent *cis*-eQTLs from AdipoXpress (n_max_=2,344)^41^, alongside fine-mapped *cis*-eQTLs and *cis-*sQTLs from GTEx v10 (n_max_=943)^42^.

Among the 282 IR lead variants or their proxies (r^2^>0.8), 109 were identified as *cis*-eQTLs and/or *cis*-sQTLs in SAT, including 19 at novel IR loci (**Supplementary Table 11**). Additional mapping in VAT using GTEx data revealed 27 unique variant-gene associations not observed in SAT, suggesting potential depot-specific regulatory effects. Extending the analysis to liver and skeletal muscle – key metabolic tissues implicated in IR –identified 7 liver- and 26 muscle-restricted *cis*-eQTLs, highlighting tissue-enriched regulatory candidates for future functional validation (**Supplementary Table 11**).

To refine the regulatory landscape in adipose tissue, we mapped IR loci to regulatory elements in mature adipocytes. Enhancer-to-gene (E2G) links were predicted using activity-by-contact (ABC) model implemented in STARE^43^, leveraging ROADMAP H3K27ac ChIP-seq data from adipose nuclei^33^ and ATAC-seq peaks from SBGS preadipocytes at days 4 and 14^34^. We identified 187 IR lead variants or proxies overlapping predicted enhancer or promoter contact regions (**Supplementary Table 12**), including regulatory links not captured by *cis*-eQTL analyses, such as for *RSPO3* and *LPL*^11^.

We next prioritized loci supported by both *cis*-eQTL (ASAT or VAT) and E2G evidence. In total, 72 IR loci linked to 119 genes replicated across both approaches, implicating them in adipose-specific gene regulation. For 114 of these genes, the predicted E2G links were further supported by chromatin interactions in mature adipocytes based on promoter-capture Hi-C data (**Supplementary Table 13**)^38-40^. These included 10 of our 70 novel IR loci: *LAMB1/LAMB1-AS1*, *AMFR*, *APOL5/APOL6*, *CDAN1*, *SACS-AS1*, *FMC1/LUC7L2*, *IRAG1*, *KCNU1*, *LIME1*, and *LONRF1* (**Table 1**). Among the 114 implicated genes, nine encode chemically tractable proteins (*CHD4, CYP27A1, FADS1, HCAR3, MAP2K7, MLX, NPC1, PLA2G6* and *TAOK2*), while nine correspond to targets of clinically validated drugs (*FGFR1, HCAR2, IGF2R, INSR, LHCGR, NISCH, NPC1L1, SV2A* and *VEGFA*) according to the Illuminating the Druggable Genome (IDG) resource (**Supplementary Table 13**)^47^.

**Table 1:** Ten novel insulin resistance loci prioritized by adipose-specific regulatory annotations.

| Lead SNP | Chr:Pos | Gene(s) linked | Cis-QTL Effect | Proxy SNP | Chr:Pos | Annotation | Chromatin state (E2G link) |
| --- | --- | --- | --- | --- | --- | --- | --- |
| BMI-decreasing IR loci |  |  |  |  |  |  |  |
| rs4727695 | 7:107614003 | LAMB1-AS1 | ↓ ASAT expression | rs10487278 | 7:107612345 | LAMB1 intron | Active enhancer (ABC/Hi-C) |
|  |  | LAMB1 | ↓ ASAT expression | rs78968481 | 7:107612784 | LAMB1 intron | Active enhancer (ABC/Hi-C) |
| BMI-neutral IR loci |  |  |  |  |  |  |  |
| rs9937484 | 16:56464517 | AMFR | ↑ ASAT & VAT expression | rs140279056 | 16:56458908 | AMFR intron | Active/Flanking TSS (ABC/Hi-C) |
| rs9610329 | 22:36042986 | APOL5 | ↓ ASAT & VAT expression | rs9610329 | 22:36042986 | Upstream APOL6 | Active enhancer (ABC/Hi-C) |
|  |  | APOL6 | ↓ ASAT & VAT expression | rs9610329 | 22:36042986 | Upstream APOL6 | Active enhancer (ABC/Hi-C) |
| rs12917018 | 15:43024063 | CDAN1 | ↓ ASAT expression | rs4265781 | 15:43028749 | CDAN1 missense | Active/Flanking TSS (ABC/Hi-C) |
|  |  |  |  | rs7167392 | 15:43028592 | CDAN1 synonymous | Active/Flanking TSS (ABC/Hi-C) |
| rs3829352 | 13:23898975 | SACS-AS1 | ↓ ASAT expression | rs34656784 | 13:23894050 | SGCG intron | Active/Flanking TSS (ABC/Hi-C) |
|  |  |  |  | rs3864968 | 13:23893798 | SGCG intron | Quiescent/Low (ABC/Hi-C) |
| rs11772771 | 7:139058803 | FMC1 | ↑ ASAT expression | rs11765421 | 7:139025094 | FMC1 Upstream | Active TSS (ABC/Hi-C) |
|  |  |  |  | rs10265 | 7:139026152 | FMC1 missense | Active TSS (ABC/Hi-C) |
|  |  |  |  | rs6947215 | 7:139026638 | FMC1 intron | Active TSS (ABC/Hi-C) |
|  |  |  |  | rs6965143 | 7:139026608 | FMC1 intron | Active TSS (ABC/Hi-C) |
|  |  | LUC7L2 | ↑ VAT expression | rs3923186 | 7:139044891 | LUC7L2 5'UTR | Active TSS (ABC/Hi-C) |
| rs4909945 | 11:10673739 | IRAG1 | Splicing ASAT & VAT | rs1544861 | 11:10679441 | IRAG1 intron | Active enhancer (ABC/Hi-C) |
| rs7816345 | 8:36846109 | KCNU1 | ↓ ASAT expression | rs56825414 | 8:36758946 | KCNU1-AS1 intron | Active enhancer/flanking TSS (ABC/Hi-C) |
| rs1151625 | 20:62369997 | LIME1 | ↓ ASAT expression | rs1291208 | 20:62330417 | ARFRP1 3'UTR Upstream | Active/flanking TSS (ABC/Hi-C) |
|  |  |  |  | rs4809331 | 20:62370890 | Upstream SLC2A4RG | Active/flanking TSS (ABC/Hi-C) |
| rs4474021 | 8:12632903 | LONRF1 | ↓ ASAT expression | rs62486442 | 8:12623463 | LINC03019 exon | Active enhancer/Quiescent low (ABC) |
|  |  |  |  | rs7464506 | 8:12624425 | LINC03019 intron | Active enhancer/Quiescent low (ABC) |

The table lists ten novel IR loci with consistent evidence for a specific effector gene, supported by both adipose tissue *cis*-eQTL signals and adipocyte E2G annotations. **Lead SNP:** Variant showing the strongest association at each locus in the multi-trait GWAS. **Chr:Pos:** Genomic coordinates (Build 37; Open Targets Genetics<sup>56</sup>). **Gene(s) linked:** Effector gene(s) supported by *cis*-eQTL in ASAT and/or VAT. ***cis*-eQTL effect:** Direction of expression or splicing QTL effect for the IR-increasing allele in ASAT or VAT. **Annotated SNP:** Lead variant or proxy in high LD ( $r^2 > 0.8$ ) with the strongest regulatory evidence relative to the linked gene. **Annotation:** Genomic context of the lead or proxy SNP relative to the linked gene. **Chromatin state:** Regulatory state in human adipocytes based on Regulome DB<sup>88,89</sup>. **E2G link:** Evidence for enhancer-to-gene links from ABC modeling and/or promoter-capture Hi-C. **ASAT:** abdominal subcutaneous adipose tissue; **VAT:** visceral adipose tissue; **QTL:** quantitative trait locus; **E2G:** enhancer-to-gene; **SNP:** single-nucleotide polymorphism; **ABC:** activity-by-contact; **TSS:** transcription start site.

Among these loci, we further experimentally characterized *LAMB1* (**Fig. 4**), where two intronic variants overlapped both ABC-predicted and promoter-capture Hi-C-validated enhancer-promoter contact regions (**Table 1**). *LAMB1* encodes the β1 subunit of laminin, a core component of the basement membrane that surrounds adipocytes and mediates cell-matrix adhesion and ECM organization^38^, processes essential for adipocyte differentiation and function. During adipogenesis, *LAMB1* expression peaks at intermediate stages while remaining low in early and mature adipocytes (**Supplementary Information**), consistent with the extensive ECM remodeling that occurs during intermediate differentiation.

**Fig. 4:**
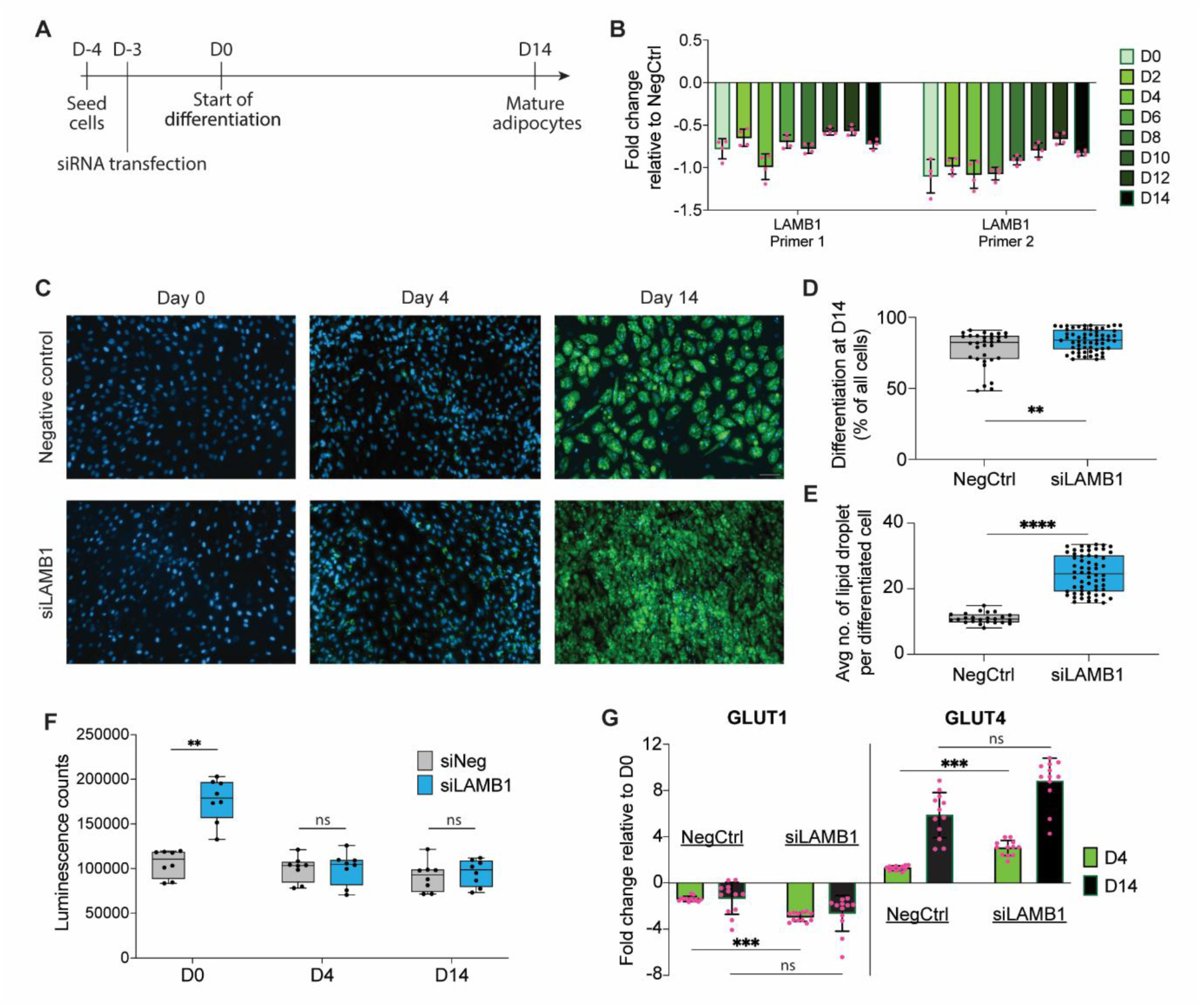
Functional characterization of *LAMB1* knockdown in SGBS cells. **A)** SGBS preadipocytes were transfected with a pool of three siRNAs targeting *LAMB1* prior to induction of differentiation and cultured to mature adipocytes. **B)** RT-qPCR quantification of *LAMB1* expression during differentiation following siRNA-mediated knockdown (siLAMB1), measured with two primer pairs targeting distinct regions of the gene. Values are normalized to negative control siRNA (NegCtrl). Data are presented as mean ± SEM. **C)** Representative fluorescence microscopy images of SGBS cells at preadipocyte (day 0), immature adipocyte (day 4) and mature adipocyte (day 14) stages following siLAMB1 or NegCtrl treatment. Lipid droplets are stained with BODIPY (green) and nuclei with Hoechst (blue). Scale bar: 100 μm. **D)** Quantification of differentiated cells at day 14 expressed as a percentage of all cells. **E)** Quantification of the average number of lipid droplets per differentiated cell at day 14 in siLAMB1 versus NegCtrl, based on BODIPY staining in panel C. **F)** Glucose uptake measured at distinct timepoints, presented as luminescence counts (relative light units) from glucose uptake assays. **G)** RT-qPCR quantification of *GLUT1* and *GLUT4* expression at days 4 and 14 of differentiation following siLAMB1 or NegCtrl treatment, relative to day 0. Data are presented as mean ± SEM.

The IR-increasing allele at the *LAMB1* locus was associated with reduced *LAMB1* expression in ASAT, suggesting that disrupted enhancer activity may contribute to adipose dysfunction and IR susceptibility (**Table 1**). Consistent with a regulatory role, *LAMB1* locus showed increased chromatin accessibility from day 4 onward in SGBS cells (**Supplementary Table 12**).

To test whether *LAMB1* regulates adipogenesis, we performed siRNA-mediated knockdown in SBGS preadipocytes. *LAMB1* silencing promoted adipogenesis, evidenced by increased adipocyte formation, greater lipid accumulation and enlarged lipid droplets (**Fig. 4, Supplementary Table 14 and Supplementary Fig. 7**). Knockdown also enhanced glucose uptake at day 0 and accelerated the transition from GLUT1- to GLUT4-mediated uptake, indicating an early metabolic shift that supports differentiations toward mature adipocyte states (**Fig. 4F-G**).

*LAMB1* silencing promoted adipogenesis, evidenced by increased adipocyte formation, greater lipid accumulation, and enlarged lipid droplets. This was accompanied by enhanced glucose uptake at day 0 and an accelerated transition from GLUT1- to GLUT4-mediated uptake, indicating an early metabolic shift that supports differentiation toward mature adipocytes (**Fig. 4F-G**).

We next examined *LAMB1* expression in adipocytes differentiated from human primary adipose mesenchymal stem cells (AMSCs) from 28 donors^48^. *LAMB1* expression correlated inversely with lipid accumulation and the number of large lipid droplets in late-stage differentiating adipocytes (day 8) (**Methods**, **Supplementary Fig. 8**), consistent with the knockdown phenotype. Under free fatty acid (FFA) exposure, *LAMB1* expression was significantly downregulated in day-14 adipocytes (**Supplementary Fig. 9A**) and was correlated with 10 imaging-derived phenotypes^48^ (FDR < 0.05), including reduced mature adipocyte morphology, decreased lipid accumulation in large droplets, and dysregulation of actin, Golgi, and plasma membrane (AGP) features (**Supplementary Table 15**, **Supplementary Information**). Genes co-varying with these *LAMB1*-associated phenotypes were enriched for pathways involved in lipid metabolism, impaired adipocyte insulin signaling (**Supplementary Table 15**) and focal adhesion and growth factor signal integration (**Supplementary Fig. 10**). Collectively, these findings indicate that *LAMB1*, through its role in ECM structure and cell-matrix adhesion, may contribute to maintaining adipocyte morphology, lipid storage, and insulin signaling – processes that become impaired under lipotoxic conditions.

### Missense variants in *PLAUR* and *INPP5A* implicate inflammation and calcium signaling in IR

To identify common coding variants contributing to IR, we took forward fine-mapped variants with posterior inclusion probability (PIP) > 0.8, suggesting a likely causal role. Fine-mapping was performed using the Causal Robust Mapping Method in Meta-Analysis (CARMA) tool^49^ and variants were annotated as missense, synonymous, or predicted loss-of-function in Ensembl’s Variant Effect Predictor (**Supplementary Table 16**)^50^. This approach identified two coding variants: Leu317Pro in *PLAUR* (rs4760-G, PIP=0.83, MAF_EUR_=0.16), encoding the urokinase-type plasminogen activator receptor (uPAR), and Lys45Arg in INPP5A (rs1133400-G, PIP=0.90, MAF_EUR_ =0.22), encoding inositol polyphosphate-5-phosphatase (I5PA) (**Fig. 5A**). Despite their relatively common allele frequencies, in silico predictions suggested these variants exert subtle but biologically meaningful effects on uPAR and IP5A function (**Fig. 5B**). Structural modeling data were only available for the *INPP5PA* Lys45Arg variant. Across five independent algorithms, predictions consistently indicated a mild positive change in Gibbs free energy (ΔΔG in kcal/mol), consistent with reduced IP5A stability (**Fig. 5B, Supplementary Information**).

**Fig. 5:**
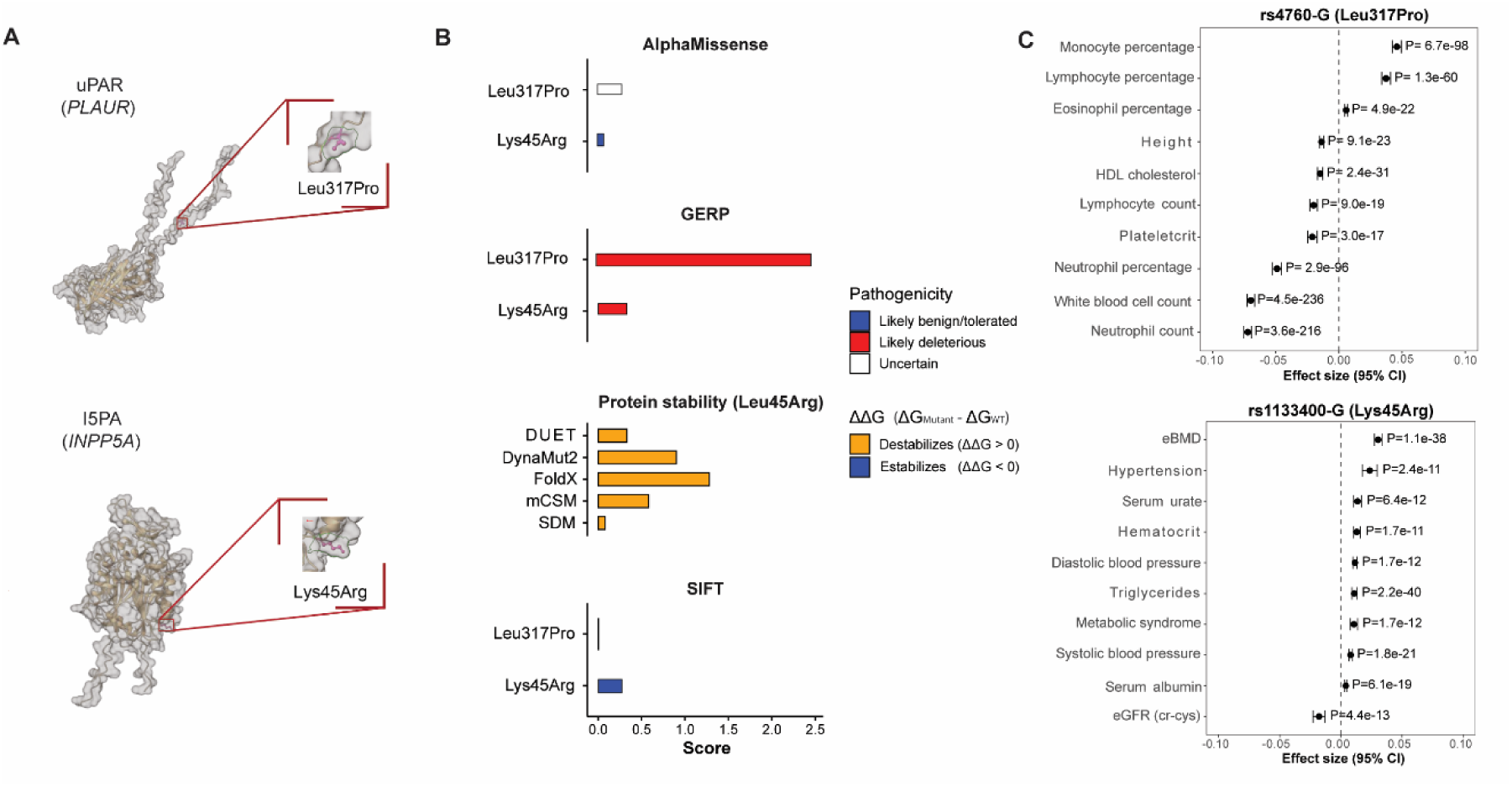
Functional characterization of Leu317Pro in *PLAUR* (encoding uPAR) and Lys45Arg in INPP5A (encoding IP5A). **A**) Structural representation of UPAR and IP5A highlighting the positions of the Leu317Pro and Lys45Arg substitutions, respectively. **B)** Pathogenicity and protein stability predictions for both variants. Pathogenicity scores were obtained from Open Platform Genetics^48^, with likely benign variants shown in blue and likely pathogenic variants in red. Protein stability predictions are reported as ΔΔG (kcal/mol), with destabilizing effects (ΔΔG > 0) shown in orange and stabilizing effects (ΔΔG < 0) in blue. **C)** Top 10 trait associations from European phenome-wide association analysis reported in Association to Function Knowledge Portal^44,45^. **eBMD**: estimated bone mineral density; **eGFR** cr-cys: estimated glomerular filtration rate computed with creatinine and cystatin C.

The *PLAUR* Leu317Pro variant has previously been associated at genome-wide significance with an IR surrogate, TG/HDL ratio^12^. Leu317Pro is the strongest genetic determinant of circulating soluble uPAR ^43^, a clinically recognized biomarker and active modulator of immune signaling, implicating uPAR-mediated inflammatory pathways in IR. Supporting this, our phenome-wide association analysis^44,45^ revealed a broad trait signature consistent with systemic inflammation **(Fig. 5C, Supplementary Table 17**).

I5PA is a key regulator of intracellular calcium signaling^38^. Phenome-wide association analysis revealed that in addition to IR, Lys45Arg is associated with several metabolic and physiological traits: heel estimated bone mineral density, TG levels, kidney function (serum creatine and cystatin C), hematocrit, hypertension, and metabolic syndrome (**Fig. 5C, Supplementary Table 17**). This pleiotropy suggests that Lys45Arg-mediated destabilization of INP5A may impair calcium signaling across multiple tissues, thereby contributing to IR and broader metabolic dysfunction.

### Mendelian randomization implicates plasma KLK1 levels as a causal mediator of IR

To investigate whether IR-associated loci exert their effects via modulation of circulating protein levels, we intersected IR lead variants and their proxies (r^2^>0.8) with fine-mapped *cis*-pQTLs reported in Open Targets Platform (n_max_= 34,557)^54,55,56^. This analysis identified 29 genome-wide significant *cis*-pQTL associations across 20 independent IR loci (**Supplementary Table 18)**. Of these, seven mapped to chemically tractable proteins (ACO3, AXL, CTSF, KLK1, KLK13, LPL, and PLAUR) and three to proteins with approved drugs (F2, IGF2R, SV2A)^47^ (**Supplementary Table 18**).

To assess causality, we performed two-sample Mendelian randomization (MR)^57^ using independent *cis*-pQTLs as instruments and FI_adjBMI_^10^ as the outcome, excluding sample overlap between exposure and outcome GWAS datasets (**Supplementary Fig. 11**). Extensive sensitivity analyses were performed to mitigate bias from horizontal pleiotropy (**Methods, Supplementary Table 18, Supplementary Fig. 12**). The MR results indicated that higher genetically predicted plasma KLK1 levels are causally associated with increased FI_adjBMI_ (P_IVW_=P=1.57x10^-6^) (**Supplementary Table 19**). KLK1 encodes the serine protease enzyme kallikrein-1 involved in bradykinin generation, a key component of the kallikrein-kinin system that regulates vascular tone, inflammation, and pain signaling^38^. Administration of recombinant kallikrein-1 has shown anti-hyperglycemic effects in rodent models^58^, supporting relevance of KLK1 as a potential mediator of IR.

## Discussion

In this study, we identified 282 genetic loci associated with IR by integrating three clinical surrogates of IR: FI_adjBMI_, HDL cholesterol, and TG. Among these, 70 represent novel loci, underscoring the power of our multi-trait approach to uncover previously undetected genetic signals. This expanded catalog of IR-associated loci provided a more nuanced view of the genetic architecture of IR, revealing both adipose-specific and systemic mechanisms and nominating new targets for mechanistic and therapeutic exploration.

Polygenic risk score analyses demonstrated that IR loci are linked to an adverse fat distribution phenotype, characterized by reduced subcutaneous fat and increased visceral and ectopic fat, without a corresponding increase in BMI. Stratifying individual loci by their direction of effect on BMI uncovered biologically distinct subgroups with differential associations across fat depots and distinct regulatory activity during adipogenesis. Specifically, BMI-decreasing variants were associated with reduced GSAT and ASAT, alongside increased VAT, liver fat, and muscle fat, whereas BMI-neutral variants were linked to reduced GSAT and increased VAT and liver fat only. These phenotypic differences were mirrored by chromatin level evidence: BMI-decreasing loci were enriched in accessible chromatin during early adipogenesis, while BMI-neutral loci were enriched in mature adipocytes. This suggests that disruptions at distinct stages of adipogenesis may differentially impair subcutaneous fat expansion and promote ectopic lipid deposition.

Using enhancer-promoter interaction mapping, we identified putative adipose-specific regulatory mechanisms for 72 IR loci. Among these, a novel locus near *LAMB1* emerged as a compelling candidate, supported by both ABC-predicted and promoter-capture Hi-C-validated enhancer-promoter interactions. Functional perturbation via siRNA-mediated knockdown of *LAMB1* in SGBS preadipocytes promoted adipogenesis, consistent with inverse correlations between *LAMB1* expression and lipid accumulation phenotypes in AMSC-derived adipocytes. However, under lipotoxic stress, lower *LAMB1* expression was associated with alteration in adipocyte morphology, reduced lipid storage, and dysregulation of actin-Golgi-plasma membrane features linked to insulin signaling.

Overall, these results suggest that *LAMB1* regulates adipocyte differentiation and structural homeostasis via ECM and cytoskeleton remodeling, offering a mechanistic explanation for IR-increasing alleles that reduce *LAMB1* expression. This interpretation is consistent with previous evidence linking *LAMB1* knockdown to ECM remodeling and insulin-related signaling in endothelial cells^59^, as well as reports of context-dependent modulation of *LAMB1* expression in adipose tissue under metabolic stress, such as increased SAT expression in high-fat diet mouse models^60^.

At the systemic level, we identified two common missense variants – Lys45Arg in *INPP5A* and Leu317 in *PLAUR* – implicating calcium signaling and inflammation, respectively, in IR pathogenesis. Phenome-wide association analyses revealed broad pleiotropic effects for both variants, consistent with their roles in systemic metabolic and immune regulation.

Integration of proteomic QTL data with MR analyses further identified circulating levels of KLK1 as a putative causal mediator of hyperinsulinemia. KLK1 encodes kallikrein-1, a serine protease involved in extracellular matrix remodeling and generation of bradykinin – a vasoactive peptide that modulates inflammation and vascular tone. The observed increase in fasting insulin levels aligns with preclinical studies showing that short-term administration of recombinant human KLK1 elevates fasting insulin while improving whole-body glucose utilization and β-cell function^58^. These findings suggest that KLK1 may exert anti-hyperglycemic effects, potentially enhancing insulin secretion independently of insulin resistance.

Our study has several limitations. First, the multi-trait GWAS was limited by the smaller sample size for FI_adjBMI_ compared with lipid traits, limiting power to detect associated loci. Second, analyses were constrained by the smaller sample sizes in non-European ancestries, limiting generalizability across populations. Third, sex-specific genetic effects on IR could not be assessed, since sex-stratified summary statistics for FI_adjBMI_ are currently unavailable.

In summary, we identified 282 loci associated with IR, including 70 novel signals, and characterized their functional impact through BMI-stratified analyses, enhancer-promoter mapping, and integrative regulatory annotation. Our findings refine the genetic architecture of IR, highlight adipose tissue as a central mediator, and nominate new candidate genes and pathways for mechanistic and therapeutic investigation.

## Methods

### Identification of IR loci through multi-trait GWAS

To identify genetic variants associated with IR, we performed multi-trait GWAS using the largest available European GWAS summary statistics for FI_adjBMI_ (n_max_=151,031), HDL cholesterol, (n_max_=1,244,580) and TG (n_max_=1,253,277) (**Supplementary Table 1**)^10, 14^. Analyses were carried out using two complementary approaches: CPASSOC**^15^** and G-SEM^16^.

For CPASSOC, we first estimated the trait correlation matrix following the original software guidelines**^15^**. We then applied the S_het_ test, optimized to detect variants with heterogenous effect directions across traits – particularly relevant for IR, where variants increase FI_adjBMI_ and TG while decreasing HDL cholesterol.

For G-SEM we implemented the common factor GWAS pipeline from the *GenomicSEM* R package (v.0.0.5)**^16^**, adapted for continuous traits. Covariance matrices were computed using the *ldsc*^70^ function, and summary statistics were formatted with *sumstats*. Association testing was performed using the *commonfactorGWAS* function, with strict genomic control to account for sample overlap between lipid traits and including only HapMap3 high-quality imputed variants^71^. This conservative restriction helped ensure that associations are driven by well-measured, common variants consistently present across all traits, mitigating the potential for spurious signals inflated by the largest sample, especially with variants with lower allele frequencies (MAF∼1%) (**Supplementary Methods**). Finally, diagonally weighted least squares (DWLS) estimation was applied, which outperformed maximum likelihood in exploratory analyses (**Supplementary Methods**).

We integrated results from both methods to define IR-associated loci (**Supplementary Fig. 1**). From CPASSOC, we retained variants reaching genome-wide significance (P<5x10^-8^) and showing IR-consistent directions of effect (FI_adjBMI_-increasing alleles associated with increased TG and decreased HDL cholesterol). From G-SEM, we selected variants with P<1x10^-6^, excluding those with significant heterogeneity (Cochran’s Q P > 5x10^-8^) (**Supplementary Table 2**). Independent loci were defined via LD-clumping using PLINK 1.9^52^ and the 1000 Genomes Phase 3 version 5 reference panel^53^ (r^2^ < 0.01, ± 1Mb window). To minimize false positives, only variants meeting the CPASSOC threshold and replicating in G-SEM with consistent IR effect directions were retained as high-confidence IR loci.

To assess generalizability across ancestries, we applied CPASSOC’s S_het_ test to GWAS summary statistics from African (AFR), East Asian (EAS), Hispanic (HIS), and South Asian (SAS) populations (n = 8,101 to 119,298)^10,14^. Replication was defined as Bonferroni-corrected significance (P<0.05/number of lead SNPs).

To identify novel signals among the 282 IR-associated loci, we performed LD-clumping between the lead IR variants and previously reported IR-associated lead variants from Lotta et al^11^, Oliveri et al., DeForest et al.^12,13^, and the MAGIC Consortium^9,10,17,18,19,20^, following the approach of Oliveri et al.^12^.

### Associations with anthropometric and cardiometabolic GWAS

We evaluated the associations between IR loci and a broad panel of anthropometric and cardiometabolic traits using publically available GWAS summary statistics (**Supplementary Table 1**)^12,23-30^. Variants with minor allele frequency (MAF) < 1% or located within the extended major histocompatibility complex region (chr6:26-34Mb) were excluded. All datasets were harmonized to GRCh37/hg19 using *liftOver***^65^**, *otargen* (v.1.0.0)^66^ and *SNPloci.Hspaiens.dbSNP155.GRCh37/38* (v 0.99.24)^67,68^.

Genetic correlations were computed using the *ldsc* function in GenomicSEM, following best practices outlined by Schoeler et al. to avoid inflation^16,69^. Polygenic risk scores were calculated using the grs.summary function from the *gtx* package (v.0.0.8)^22^, based on G-SEM effect sizes. Outcome summary statistics were aligned to the IR-increasing allele using *harmonise_data* function from *TwoSampleMR* (v.0.5.7)^57^ or *snp_match* from *bigsnpr* (v1.12.18)^70^. Missing variants were replaced with high-LD proxies (r^2^>0.8) identified via *Haploreg* (v.4.2.0)^71^.

### Enrichment analyses

#### Tissue and gene-set enrichment

We performed tissue and gene-set enrichment analyses using DEPICT (v1 “rel194”) with default settings^31^. Gene-set network analyses applied a Pearson correlation cut-off of 0.3, and significance was defined as FDR < 0.05.

#### Regulatory element enrichment

We assessed enrichment of IR loci in active regulatory elements across 127 epigenomes from ROADMAP’s 15-core chromatin state model (ChIP-seq data)^33^ using GoShifter (v0.3; 10.000 permutations)^32^. Active promoter/enhancer states included: 1_TssA, 2_TSSAFlnk, 6_EnhG, 7_Enh, 10_TssBiv, and 11_BivFlnk 12_EnhBiv.^72^ Enrichment in adipose tissue was compared with consensus ATAC-seq peaks from METSIM bulk adipose tissue (n=11) and the top 100,000 peaks from SGBS preadipocytes^35^ at day 0 (predipocytes), day 4 (immature adipocytes), and 14 (mature adipocytes)^34^. Significance was defined as P < 1x10^-4^, as recommended by the authors^32^.

We further tested enrichment in SGBS ATAC-seq peaks using CHEERS (v2019)^36^. Batch effects were corrected using *ComBat_seq* from the *sva* R package (3.35.2)^73^. Bonferroni-corrected significance was set at P < 2x10^-4^ (P=0.05/25 tests).

#### Transcription factor enrichment

Adipocyte-specific transcription factor (TF) binding enrichment was assessed using ChIP-atlas^37^, and TF motif enrichment was evaluated using HOMER (v4.11.1)^39^. Enrichment was tested by comparing IR-overlapping SGBS ATAC-seq peaks to background peaks. ChIP-Atlas queries (last accessed 05/09/2025) were restricted to adipocyte-relevant cell types and build 37 (hg19). HOMER results were reported for known TF motifs only.

### Gene prioritization using *cis*-QTLs

To link IR variants to putative effector genes, we integrated *cis*-expression quantitative trait loci (*cis*-eQTLs) and *cis*-splicing QTL (*cis*-sQTLs) data from insulin-sensitive tissues: subcutaneous and visceral adipose tissue (SAT, VAT), liver, and skeletal muscle. An IR variant was considered linked to a gene if it, or a high-LD proxy (r^2^>0.8), was a conditionally independent QTL (P < 1x10^-6^) or part of a fine-mapped credible set. For SAT, we used conditionally independent *cis*-eQTLs from AdipoXpress (n_max_=2,344)^41^ and fine-mapped *cis*-eQTLs and *cis*-sQTLs from GTEx v10 (n_max_=943)^42^. For VAT, liver, and skeletal muscle, we utilized GTEx v10 fine-mapped data.

### Enhancer-to-gene strategy

To systematically link IR loci to regulatory elements modulating gene expression in adipocytes, we implemented an enhancer-to-gene (E2G) strategy in two steps.

Step 1: Regulatory elements were mapped to gene promoters in adipocytes using the STARE-adapted activity-by-contact (ABC) model^43^. Specifically, we applied the ABC_pp_ method with default parameters, integrating epigenomic data and a power-law distance function to predict enhancer-promoter contacts^43^. ABC_pp_ was run using two sources of adipocyte data: (i) ROADMAP ChIP-seq narrowPeak H3K27ac data for adipose nuclei (E063)^33^ and (ii) the top 100.000 chromatin accessibility peaks in SGBS cells at day 4 and day 14^34^.

Step 2: We assessed whether IR variants within these regulatory elements were associated with differential expression (*cis*-eQTLs) or alternative splicing (*cis*-sQTLs) of the linked genes. Enhancer-promoter and variant-gene links were reported (**Supplementary Table 12**), with emphasis on those replicated by promoter-capture Hi-C data from mature adipocytes (GSM3004355, GSE129574)^44,45^ and differentiated SGBS cells (GSE262496)^46^ (**Supplementary Table 13**).

### Identification and analysis of common coding missense variants

To identify common (MAF > 1%) coding IR variants, we performed fine-mapping with CARMA^49^ across ±500kb regions flanking each independent IR variant, using summary statistics from the common factor GWAS and the European 1000 Genomes Phase 3 version 5 reference panel (N = 503)^64^.

Posterior inclusion probabilities (PIPs) were extracted for all independent IR variants and their proxies (r^2^ > 0.8), prioritizing those with PIP > 0.8 as likely single-variant drivers. Variant annotation was performed using the *variantInfo* function from the *otargen* R package (v.1.0.0)^66^, which retrieves Variant Effect Predictor (VEP)^50^ data from the Open Targets Platform^56^. We retained missense, synonymous, and predicted loss-of-function (pLOF) variants.

Pathogenicity of coding variants was assessed using AlphaMissense^74^, SIFT^75^, GERP^76^, and FoldX^77^ scores via Open Targets Platform^56^. FoldX protein stability predictions were validated with four additional algorithms: mCSM^78^, SDM^79^, DUET^80^ and DynaMut2^81^. Protein structures were visualized with ChimeraX^82^ using AlphaFold2-predicted models^83^. Functional effects were further explored by querying phenome-wide association results (PheWAS) in the Association-to-Function (A2F) database^52,53^.

### Mendelian randomization analyses with protein quantitative trait loci

To identify proteins causally associated with IR, we annotated IR loci as putative causal *cis*-pQTLs using fine-mapped European-ancestry credible sets from INTERVAL (n_max_= 3,301) and UK Biobank Pharma Proteomics Project (UKBB-PPP-; n_max_= 34,557)^54,55^, as available in the Open Targets Platform^56^.

We then performed two-sample Mendelian randomization (MR) using the *TwoSampleMR* package (v0.5.7.), following STROBE-MR guidelines^57,84^. *Cis*-pQTL summary statistics from UKBB-PPP were used as exposures, and FI_adjBMI_ (the only hallmark IR trait without sample overlap) as the outcome^10^. To avoid violating MR assumptions^57^ (**Supplementary Fig. 11**) and minimize bias: (i) Instruments were restricted to the variant with the highest PIP from each fine-mapped *cis*-pQTL credible set, with F-statistic > 10; (ii) Only proteins with > 3 independent instruments were tested, enabling use of MR methods robust to horizontal pleiotropy (MR Egger, weighted median, weighted mode); (iii) Steiger filtering was employed to exclude variants more strongly associated with the outcome than the exposure; (iv) Sensitivity analyses, including Cochran’s Q and I^2^ heterogeneity tests, Rucker’s test, and Egger’s intercept test were applied; and (v) Pleiotropic variants were removed based on leave-one-out, funnel, and RadialMR plots^85^.

### Cell culture

SGBS preadipocytes^35,86^, provided by Dr. Martin Wabitsch (University of Ulm), were cultured at 37°C, 5% CO_2_, with media replaced every 2 days. Standard culture medium consisted of DMEM/F-12 (ThermoScientific) supplemented with 33 μM biotin, 17 μM pantothenic acid, 10% FBS, 100 IU/mL penicillin, and 100 μg/mL streptomycin. Differentiation medium was serum-free standard medium further supplemented with 0.01 mg/mL human transferrin, 100 nM cortisol, 200 pM triiodothyronine, 20 nM human insulin, 25 nM dexamethasone, 250 μM IBMX, and 2 μM rosiglitazone. On day 4, media was replaced with serum-free standard medium supplemented with 0.01 mg/mL human transferrin, 100 nM cortisol, 200 pM triiodothyronine, and 20 nM human insulin. Cells were differentiated for 14 days, plated at high density to ensure optimal differentiation capacity^35^.

### Quantitative PCR

Total RNA was isolated using the RNeasy Mini Kit (QIAGEN) according to the manufacturer’s instructions. Reverse transcription was carried out with the iScript cDNA Synthesis Kit (Bio-Rad). Gene expression was quantified on a Bio-Rad CFX384 thermal cycler using PrecisionPlus qPCR Master Mix (Primer Design) under standard cycling conditions. Relative gene expression was calculated using the ΔΔC_T_ method, normalized to *RPL13A* housekeeping gene expression. *GAPDH* served as a positive control, and *ADIPOQ* as a differentiation marker. Primer sequences are listed in **Supplementary Table 14**.

### siRNA-mediated gene knockdown

MISSION siRNA Universal Negative Control #1 and siRNAs targeting *GAPDH* and *LAMB1* (Sigma) were transfected into SGBS cells using Lipofectamine RNAiMAX (Invitrogen) according to the manufacturer’s protocol. A pool of three siRNAs targeting *LAMB1* was used. Cells were incubated with siRNA for 72 h prior to initiation of differentiation.

### Fluorescence imaging

Cells were fixed with 4% paraformaldehyde in PBS for 20 min at room temperature, washed twice with PBS, and stained with Hoechst (ThermoScientific) and BODIPY (Invitrogen) in PBS for 30 min at room temperature in the dark. Cells were kept in PBS at 4°C before imaging, performed on a Zeiss Axio Observer microscope with a 10x air objective. Fiji software was used for image analysis, with StarDist applied for nuclear and lipid droplet segmentation.

### Glucose uptake assay

Glucose uptake was measured using the Glucose Uptake-Glo kit (Promega) according to the manufacturer’s instructions. Cells were treated with siRNAs at days 0, 4, and 14 of differentiation, with incubation for 72 h prior to assay.

### *LAMB1* phenotypic and transcriptomic consequences in human AMSCs

Primary subcutaneous adipose mesenchymal stem cells (AMSCs) were obtained from 28 adult donors under informed consent and institutional approval, and isolated, expanded, and differentiated as previously described.^48^ AMSCs were induced to adipogenic differentiation (day 0, 3, 8 and 14) following established protocols^48^. At each time point, bulk RNA-seq and image-based phenotyping (LipocyteProfiler) were performed, as previously described^48^. To model lipotoxic stress, AMSCs were cultured with or without FFA stimulation during maturation.

Associations between *LAMB1* expression and LipocyteProfiler-derived cellular phenotypes were quantified using linear mixed models adjusting for age, sex, BMI, T2D and batch. P values were FDR-adjusted across features using Benjamini-Hocherg, with FDR<0.05 considered significant. To interpret significant *LAMB1*-phenotype associations, we identified co-varying gene sets by correlating each significant LipocyteProlifer feature with transcriptome-wide gene expression across samples. Pathway enrichment analysis was performed using gprofiler2 v0.2.3^87^ considering pathways at FDR<0.05 as enriched.

## Supporting information

MR-STROBE guidelines checklist for Mendelian Randomization analyses

Supplementary methods, results and figures

## Code and Data availability

The code utilized to replicate these analyses can be found in: https://github.com/MarioGuCBMR/IR_multiGWAS/tree/main. Multi-trait common factor GWAS summary statistics for IR computed with G-SEM can be found in: 10.5281/zenodo.18862458

## Acknowledgements

This work was supported by the Novo Nordisk Foundation (NNF21SA0072102, NNF22OC0074128, NNF23SA0084103) and by the National Institutes of Health (UM1DK126185, P30DK040561). Novo Nordisk Foundation Center for Basic Metabolic Research (https://cbmr.ku.dk) is an independent research center at the University of Copenhagen, partially funded by an unrestricted donation from the Novo Nordisk Foundation. MW received funding by the Federal Ministry of Research, Technology and Space (Bundesministerium für Forschung, Technologie und Raumfahrt, BMFTR) as part of the German Center for Child and Adolescent Health (DZKJ) under the funding code 01GL2407A.

