## Supplementary material for "Genome-Wide Discovery Reveals Adipose-Specific and Systemic Regulators of Insulin Resistance": MR-STROBE guidelines checklist for Mendelian Randomization analyses

**STROBE-MR checklist of recommended items to address in reports of Mendelian randomization studies**^1^ ^2^

| **Item No.** | **Section** | **Checklist item** | **Page No.** | **Relevant text from manuscript** |
| --- | --- | --- | --- | --- |
| 1 | **TITLE and ABSTRACT** | Indicate Mendelian randomization (MR) as the study’s design in the title and/or the abstract if that is a main purpose of the study | 1 | Mendelian Randomization (MR) analyses are not the main study design of this project, but a prioritization to link genetically increased blood-circulating protein levels to insulin resistance. |
|  | **INTRODUCTION** |  |  |  |
| 2 | **Background** | Explain the scientific background and rationale for the reported study. What is the exposure? Is a potential causal relationship between exposure and outcome plausible? Justify why MR is a helpful method to address the study question |  | Multi-trait genetic studies have identified IR-associated loci by leveraging genome-wide association studies (GWAS) from hallmark IR traits. In this study we have identified 282 IR-associated loci, 70 of them novel, by leveraging GWAS of fasting insulin adjusted for body mass index (FIadjBMI), high density lipoprotein (HDL) cholesterol and triglycerides (TG) with two complementary methods: cross-phenotype associations (CPASSOC) and genomic structural equation modelling (G-SEM).  To prioritize potential drug targets to treat IR, we leveraged fine-mapped protein quantitative trait loci (pQTLs) and identified that 20 IR loci are associated with differential circulating levels of up to 29 proteins, 7 of them novel. However, while these analyses link IR loci to circulating proteins, it remains unclear whether variation in the levels of these proteins contribute directly to IR-associated phenotypes.  In this study, we used Mendelian randomization (MR) as an additional prioritization approach. Specifically, we aimed to identify proteins whose genetically predicted circulating levels (exposure) influence one of the hallmarks of insulin resistance: hyperinsulinemia, measured using FIadjBMI (outcome).  The MR framework is useful in this context because, by leveraging genetic variants identified through genome-wide association studies (GWAS) as instrumental variables, it allows testing whether circulating protein levels have a potential causal effect on FIadjBMI while reducing bias from reverse causation and confounding. |
| 3 | **Objectives** | State specific objectives clearly, including pre-specified causal hypotheses (if any). State that MR is a method that, under specific assumptions, intends to estimate causal effects |  | The aim of the study is to identify novel IR loci and uncover the biology underlying the associations. In this context, the objective of the MR analysis is to further prioritize proteins that may represent potential drug targets for IR.  Specifically, we aim to prioritize proteins that might have a direct on fasting insulin levels. If pleiotropy is properly controlled, the MR framework will produce causal estimates between genetically-increased protein levels and fasting insulin. These should be interpreted with caution, since protein levels can be also affected by trans-regulation. These analyses barely serve as a prioritization method to link cis-regulation of protein expression to fasting insulin levels. |
|  | **METHODS** |  |  |  |
| 4 | **Study design and data sources** | Present key elements of the study design early in the article. Consider including a table listing sources of data for all phases of the study. For each data source contributing to the analysis, describe the following: | 5-7 |  |
|  | a) | Setting: Describe the study design and the underlying population, if possible. Describe the setting, locations, and relevant dates, including periods of recruitment, exposure, follow-up, and data collection, when available. |  | We utilized publicly available GWAS for pQTLs and FIadjBMI. Information on the recruitment of individuals and data collection can be found in the respective papers. Access to the publications can be found in Supplementary Table 1 of this manuscript. |
|  | b) | Participants: Give the eligibility criteria, and the sources and methods of selection of participants. Report the sample size, and whether any power or sample size calculations were carried out prior to the main analysis | 7–9 | For exposure, we utilized GWAS summary statistics from European ancestry for pQTL levels of 29 different proteins (n=34,557) from UK Biobank and for FIadjBMI (n=151, 031) from MAGIC Consortium. There is no sample overlap between studies. No power calculations were computed prior to analyses. |
|  | c) | Describe measurement, quality control and selection of genetic variants | 9-11 | Independent cis-pQTLs were selected for each protein if they were reported in fine-mapped pQTL credible sets (95% confidence).  Instruments were restricted to the variant with the highest PIP from each fine-mapped cis-pQTL credible set, with F-statistic > 10. |
|  | d) | For each exposure, outcome, and other relevant variables, describe methods of assessment and diagnostic criteria for diseases | 11-13 | For exposure, protein levels were measured with antibody-based technology: Olink.Units are in Normalized Protein eXpresion (NPX). GWAS were computed after NPX’s inverse-rank normalization adjusting for age, age squared, sex, age-sex interaction and age-squared-sex interaction, batch, UKB centre, UKB genetic array, time between blood sampling and measurement and 20 principal components.  Outcome: FIadjBMI GWAS is a meta-analysis. FI levels were measured in natural log-transformed pmol/L. GWAS were computed without inverse-rank normalization and adjusting for BMI. Other co-variates varied according to the study, though in most cases included age, age-squared and sex.  In both cases, individuals with T2D were excluded. |
|  | e) | Provide details of ethics committee approval and participant informed consent, if relevant | 9 | We utilized publicly available GWAS. |
| 5 | **Assumptions** | Explicitly state the three core IV assumptions for the main analysis (relevance, independence and exclusion restriction) as well assumptions for any additional or sensitivity analysis | 11–12 | We ensured IV relevance by:   1) Selecting independent cis-pQTL from fine-mapped credible sets (95% confidence). 2) Instruments were restricted to the variant with the highest PIP from each fine-mapped cis-pQTL credible set, with F-statistic > 10.   We ensure independence from the outcome by:   1) Only instrumenting variants that passed steiger filtering (i.e. more correlated with exposure than outcome). 2) Removed outliers detected by a combination of Rucker test and RadialMR. 3) Performed sensitivity tests and diagnostic analyses to assess pleiotropy and heterogeneity.   We ensured we did not violate the exclusion restriction assumption by:   1) Removed outliers detected by a combination of Rucker test and RadialMR. 2) Performed sensitivity tests and diagnostic analyses to assess pleiotropy and heterogeneity.   Importantly: we only performed MR analyses for those proteins with, at least, 3 independent cis-pQTL signals. This allowed to only including analyses were methods that control for pleiotropy (MR-Egger, weighted median and weighted mode) could be performed. |
| 6 | **Statistical methods: main analysis** | Describe statistical methods and statistics used |  |  |
|  | a) | Describe how quantitative variables were handled in the analyses (i.e., scale, units, model) | 12-13 | GWAS for protein levels were in inverse-ranked normalized NPX units. GWAS for FIadjBMI were in natural log-transformed pmol/L. |
|  | b) | Describe how genetic variants were handled in the analyses and, if applicable, how their weights were selected | 11-12 | For each protein, we selected the cis-pQTL with the largest PIP across independent credible sets (95% confidence). |
|  | c) | Describe the MR estimator (e.g. two-stage least squares, Wald ratio) and related statistics. Detail the included covariates and, in case of two-sample MR, whether the same covariate set was used for adjustment in the two samples |  | We utilized 4 MR methods: inverse variance weighted, MR-Egger, weighted median and weighted mode. All of these methods are based on meta-analysing independent wald ratios.  Covariates for exposure and outcome are described in section 4D. Importantly, only the outcome GWAS was adjusted for BMI.  There is no sample overlap across exposure and outcome studies. |
|  | d) | Explain how missing data were addressed |  | For a missing IV in the outcome GWAS, we assessed whether any of its LD proxies (r2>0.8) could be utilized instead. Proxies were retrieved from HaploReg. |
|  | e) | If applicable, indicate how multiple testing was addressed |  | NA. |
| 7 | **Assessment of assumptions** | Describe any methods or prior knowledge used to assess the assumptions or justify their validity |  | This is an exploratory analyses. |
| 8 | **Sensitivity analyses and additional analyses** | Describe any sensitivity analyses or additional analyses performed (e.g. comparison of effect estimates from different approaches, independent replication, bias analytic techniques, validation of instruments, simulations) | 12-15 | No additional analyses were performed aside from the sensitivity tests used to account for pleiotropy. |
| 9 | **Software and pre-registration** |  |  |  |
|  | a) | Name statistical software and package(s), including version and settings used | 11-12 | *TwoSampleMR* (v0.5.7) and *RadalMR* (v1.1.0) |
|  | b) | State whether the study protocol and details were pre-registered (as well as when and where) |  | This study was not pre-registered. |
|  | **RESULTS** |  |  |  |
| 10 | **Descriptive data** |  |  |  |
|  | a) | Report the numbers of individuals at each stage of included studies and reasons for exclusion. Consider use of a flow diagram | 7-9 | For exposure, we utilized GWAS summary statistics from European ancestry for pQTL levels of 29 different proteins (n=34,557) from UK Biobank and for FIadjBMI (n=151, 031) from MAGIC Consortium. There is no sample overlap between studies. Individuals with T2D were excluded for FIadjBMI. More information on exclusion criteria can be found in the original publications, which can be accessed from Supplementary Table 1. |
|  | b) | Report summary statistics for phenotypic exposure(s), outcome(s), and other relevant variables (e.g. means, SDs, proportions) |  | Summary statistics are publicly available and can be obtained in their respective publications. Links to download the data can be accessed from Supplementary Table 1. |
|  | c) | If the data sources include meta-analyses of previous studies, provide the assessments of heterogeneity across these studies |  | For FIadjBMI, any variant that presented P<1x10-05 for heterogeneity test was reviewed by, at least 2 analyst. Other QC assessments of heterogeneity across studies can be found in the Supplementary Information of Chen et al 2021. Particularly in Single-ancestry and trans-ancestry meta-analyses section b: manual curation of single-ancestry index and lead variants and trans-ancestry lead variants. |
|  | d) | For two-sample MR:  i.  Provide justification of the similarity of the genetic variant-exposure associations between the exposure and outcome samples  ii.  Provide information on the number of individuals who overlap between the exposure and outcome studies |  | GWAS were performed in individuals from European ancestry. There is no sample overlap between studies. |
| 11 | **Main results** |  |  |  |
|  | a) | Report the associations between genetic variant and exposure, and between genetic variant and outcome, preferably on an interpretable scale |  | All intermediary files utilized to compute MR estimates can be found in the github for the project: https://github.com/MarioGuCBMR/IR_multiGWAS/tree/main |
|  | b) | Report MR estimates of the relationship between exposure and outcome, and the measures of uncertainty from the MR analysis, on an interpretable scale, such as odds ratio or relative risk per SD difference |  | Results for all tested pQTL–FIadjBMI associations are reported in Supplementary Table 19. Here we highlight the most consistent finding across method: genetically increased KLK1 levels were associated with higher FIadjBMI. The inverse-variance weighted (IVW) analysis estimated an effect of 0.0083 (95% CI: 0.0049–0.0117). This result indicates that a 1 SD genetically predicted increase in circulating KLK1 levels (measured in inverse-rank normalized NPX units) is associated with a 0.0082 increase in FIadjBMI (log-transformed pmol/L). |
|  | c) | If relevant, consider translating estimates of relative risk into absolute risk for a meaningful time period |  | Not relevant in this particular case. |
|  | d) | Consider plots to visualize results (e.g. forest plot, scatterplot of associations between genetic variants and outcome versus between genetic variants and exposure) |  | Scatter plots, leave-one-out plots and funnel plots for all analyses can be found in the github: <https://github.com/MarioGuCBMR/IR_multiGWAS/tree/main>. For KLK1 these plots have been reported in Supplementary Figure 12. |
| 12 | **Assessment of assumptions** |  |  |  |
|  | a) | Report the assessment of the validity of the assumptions | 11-12,15-17 | We conducted a comprehensive set of sensitivity analyses to evaluate potential violations of the independence and exclusion restriction assumptions. Horizontal pleiotropy was assessed using the MR-Egger intercept test and the Rücker framework, which also enabled comparison of model fit between inverse-variance weighted (IVW) and MR-Egger approaches. To further identify influential variants and potential sources of pleiotropy, we performed leave-one-out analyses and inspected funnel plots.  We subsequently applied RadialMR to systematically detect and remove outlier variants. Following outlier exclusion, all primary and sensitivity analyses were repeated to determine whether the observed estimates were robust to pleiotropic effects. |
|  | b) | Report any additional statistics (e.g., assessments of heterogeneity across genetic variants, such as *I^2^*, Q statistic or E-value) |  | We report Cochran’s Q and I^2^. |
| 13 | **Sensitivity analyses and additional analyses** |  |  |  |
|  | a) | Report any sensitivity analyses to assess the robustness of the main results to violations of the assumptions | 15-20 | RadialMR was applied irrespective of evidence for horizontal pleiotropy from the MR-Egger intercept. In the majority of pQTL–FIadjBM analyses where no pleiotropic bias was detected, RadialMR did not identify outlier variants, further supporting the robustness of the findings to potential violations of instrumental variable assumptions. |
|  | b) | Report results from other sensitivity analyses or additional analyses |  | No other analyses were performed. |
|  | c) | Report any assessment of direction of causal relationship (e.g., bidirectional MR) |  | We did not perform bi-directional MR. |
|  | d) | When relevant, report and compare with estimates from non-MR analyses |  | NA. |
|  | e) | Consider additional plots to visualize results (e.g., leave-one-out analyses) |  | Additional plots can be found in Supplementary Figure 12 and in the github for this project. |
|  | **DISCUSSION** |  |  |  |
| 14 | **Key results** | Summarize key results with reference to study objectives |  | Genetically increased KLK1 blood levels are associated with increased FIadjBMI. With this result, we have prioritized a potential drug target for IR treatment. |
| 15 | **Limitations** | Discuss limitations of the study, taking into account the validity of the IV assumptions, other sources of potential bias, and imprecision. Discuss both direction and magnitude of any potential bias and any efforts to address them |  | Our study design focused on FIadjBMI to prioritize proteins with potential direct effects on hyperinsulinemia. Inclusion of additional insulin resistance–related traits may broaden the set of prioritized targets. Furthermore, all analyses were restricted to individuals of European ancestry; replication in diverse populations will be important to validate and extend these findings, particularly with respect to KLK1 as a potential therapeutic target. |
| 16 | **Interpretation** |  |  |  |
|  | a) | Meaning: Give a cautious overall interpretation of results in the context of their limitations and in comparison with other studies |  | Our exploratory analyses prioritized KLK1 as a potential drug target for IR and diabetes. Indeed, we identified that genetically-increased KLK1 levels are associated with increased levels of fasting insulin. This is consistent with results of pre-clinical studies showing that administration of KLK1 elevates fasting insulin levels and improves beta-cell function (Kolodka et al. 2014). |
|  | b) | Mechanism: Discuss underlying biological mechanisms that could drive a potential causal relationship between the investigated exposure and the outcome, and whether the gene-environment equivalence assumption is reasonable. Use causal language carefully, clarifying that IV estimates may provide causal effects only under certain assumptions |  | Taking into account pre-clinical data from Kolodka et al. 2014, a potential causal mechanism underlying the associations between KLK1 and hyperinsulinemia might be the effect of KLK1 in the pancreas. Potentially, effects of KLK1 in the pancreas can lead to increase insulin secretion. This would explain the increase of insulin and decrease in glycemia reported in Kolodka et al.  Additionally, KLK1 levels in peripheral tissues can be translated into increased insulin sensitivity, which would explain why fasting insulin is increased yet hyperglycemia is decreased in the results reported by Kolodka et al.  Notably, KLK1 is an in-situ serine protease which cleaves kinogens to bradykinin, which acts as a vasodilator. |
|  | c) | Clinical relevance: Discuss whether the results have clinical or public policy relevance, and to what extent they inform effect sizes of possible interventions |  | These findings have limited direct clinical applicability, as the study was designed to prioritize potential therapeutic targets. However, if KLK1 levels predispose to or correlate with insulin resistance, they may have utility as a biomarker for its detection in clinical settings. This interpretation remains speculative and serves primarily to illustrate the potential clincial relevance of our results. |
| 17 | **Generalizability** | Discuss the generalizability of the study results (a) to other populations, (b) across other exposure periods/timings, and (c) across other levels of exposure |  | Generalizability to other populations warrants further investigation, although this lies beyond the scope of the present secondary analysis, which was designed to prioritize proteins linked to IR loci by leveraging cis-pQTL data. |
|  | **OTHER INFORMATION** |  |  |  |
| 18 | **Funding** | Describe sources of funding and the role of funders in the present study and, if applicable, sources of funding for the databases and original study or studies on which the present study is based |  | This work was supported by the Novo Nordisk Foundation (NNF21SA0072102, NNF22OC0074128, NNF23SA0084103) and by the National Institutes of Health (UM1DK126185, P30DK040561). Novo Nordisk Foundation Center for Basic Metabolic Research (https://cbmr.ku.dk) is an independent research center at the University of Copenhagen, partially funded by an unrestricted donation from the Novo Nordisk Foundation. MW received funding by the Federal Ministry of Research, Technology and Space (Bundesministerium für Forschung, Technologie und Raumfahrt, BMFTR) as part of the German Center for Child and Adolescent Health (DZKJ) under the funding code 01GL2407A. |
| 19 | **Data and data sharing** | Provide the data used to perform all analyses or report where and how the data can be accessed, and reference these sources in the article. Provide the statistical code needed to reproduce the results in the article, or report whether the code is publicly accessible and if so, where |  | *Scatter plots, leave-one-out plots and funnel plots for all analyses can be found in the github: https://github.com/MarioGuCBMR/IR_multiGWAS/tree/main. For KLK1 these plots have been reported in Supplementary Figure 12.* |
| 20 | **Conflicts of Interest** | All authors should declare all potential conflicts of interest |  | M .C. has received consulting honoraria from Novo Nordisk, Genentech, Pfizer; is part of the SAB of Nestle, SixPeaks Bio, Waypoint Bio, Verdiva Bio, and served as a scientific co-founder of Sidera Bio in which she holds equity. |

This checklist is copyrighted by the Equator Network under the Creative Commons Attribution 3.0 Unported (CC BY 3.0) license.

1.	Skrivankova VW, Richmond RC, Woolf BAR, Yarmolinsky J, Davies NM, Swanson SA, et al. Strengthening the Reporting of Observational Studies in Epidemiology using Mendelian Randomization (STROBE-MR) Statement. JAMA. 2021;under review.

2.	Skrivankova VW, Richmond RC, Woolf BAR, Davies NM, Swanson SA, VanderWeele TJ, et al. Strengthening the Reporting of Observational Studies in Epidemiology using Mendelian Randomisation (STROBE-MR): Explanation and Elaboration. BMJ. 2021;375:n2233.
