## Supplementary methods, results and figures for "Genome-Wide Discovery Reveals Adipose-Specific and Systemic Regulators of Insulin Resistance"

### Supplementary Information

#### Multi-trait GWAS modeling and genomic control

In the main text we highlighted the complementary rationale for applying both CPASSOC[1] and GenomicSEM (G-SEM)[2] to test multi-trait associations with insulin resistance. CPASSOC ($S_{het}$ test) is optimized to detect directionally heterogeneous multi-trait effects (e.g. $FI_{adjBMI}$-increasing alleles associated with TG increase and HDL decrease) whereas G-SEM models shared genetic architecture across traits and provides effect size estimates suitable for downstream analyses. Here we provide technical details on model performance, genomic control, and the criteria used to define high-confidence IR loci.

CPASSOC ($S_{het}$ test) identified 80,285 genome-wide significant multi-trait associations with IR (**Supplementary Fig. 1**). To ensure robustness of our association signals, as well as to provide multi-trait effect sizes and standard errors that are not available from CPASSOC, we performed a common factor GWAS using G-SEM. Two G-SEM estimation methods were tested: diagonal weighted least squares (DWLS) and maximum likelihood (ML). DWLS identified 3,815 genome-wide significant associations with a predicted sample size of 3,981,077 (based on Mallard et al.[3]), whereas ML identified no genome-wide significant associations and yielded a predictive effective sample size of 62,114. Given the underperformance of ML, we proceeded with the DWLS results.

The large effective sample size estimated for DWLS raised concerns about potential inflation in connection with the uneven and overlapping sample sizes between the GWAS traits analyzed. However, in accordance with strict genomic control applied during G-SEM computation, QQ plot for the DWLS multi-trait results showed no evidence of inflation (**Supplementary Fig. 2**) and behaved similarly to that of recently published TG/HDL GWAS summary statistics[4] ($\lambda_{TG/HDL} = 1.32$; $\lambda_{IR} = 1.47$). This indicates that strict genomic control effectively compensates for uneven and overlapping sample sizes.

Notably, DWLS identified fewer genome-wide significant variants compared with CPASSOC, suggesting reduced sensitivity. We believe the reason behind these results is a combination of the strict genomic control setting plus computing G-SEM multivariate association only with HapMap3[5] high-quality imputed variants. This might limit loci discovery, but boosts robustness, particularly in a setting with uneven sample sizes between $FI_{adjBMI}$ ($n_{max}$=151,031)[6], HDL cholesterol, ($n_{max}$=1,244,580)[7] and TG ($n_{max}$=1,253,277)[7]. Thus, to balance specificity and sensitivity in insulin resistance variant discovery, we retained only variants that replicated across both CPASSOC and G-SEM (**Supplementary Fig. 1**), applying a more permissive threshold ($P<1x10^{-6}$) for G-SEM associations than for CPASSOC ($P<5x10^{-8}$). This combined strategy ensured robust identification of IR loci while minimizing potential for false positives arising from either method.

#### Sensitivity analyses to evaluate the robustness of multi-trait associations

##### Genetic correlations

We estimated genetic correlations between the common factor GWAS and $FI_{adjBMI}$[6], HDL cholesterol[7], and TG[7], five glycemic traits related to insulin/glucose biology[6,8], six IR-related diseases[9], and seven anthropometric traits[10-12] (**Supplementary Table 1, Supplementary Fig. 3**). In accordance with expectations, the common factor GWAS demonstrated strong genetic correlations[13] with $FI_{adjBMI}$ ($r_g$ = 0.47 [0.39, 0.55], P =$9.90x10^{-31}$), HDL cholesterol ($r_g$ = -0.85 [-0.92,-0.77], P=$1.40x10^{-105}$), and TG ($r_g$ =0.84 [0.80, 0.93], P=$2.20x10^{-85}$), as with other markers of insulin resistance, such as TG/HDL ratio[4] ($r_{g-TG/HDL}$=0.93 [0.85, 1.00], P=$1.20x10^{-105}$) and BMI-adjusted Stumvoll insulin sensitivity index[8] ($ISI_{adjBMI}$, $r_g$=-0.35 [-0.43,-0.27] , P=$2.10x10^{-18}$), and the risk of type 2 diabetes[9] ($r_g$ = 0.56 [0.51, 0.61], P= $1.60x10^{-89}$). This confirms that the common factor GWAS multi-trait associations are well aligned with traits and diseases known to be mechanistically linked to insulin resistance.

##### Replication of known IR loci

We next assessed whether the common factor GWAS recapitulated previously reported IR loci. Comparing the 282 IR loci, including lead variants and all variants with $r^2$ ≥0.01 within ±1Mb from the lead variant[14,15] against the lead variants reported by Lotta et al., Oliveri et al., DeForest et al., and the MAGIC consortium[,16-22,] , revealed that 212 (75%) independent associations overlapped with known IR loci (**Supplementary Table 2**). A Manhattan plot of the 282 IR variants (**Supplementary Fig. 4**) further highlights that the majority of the most significant loci corresponds to 53 IR loci previously reported by Lotta et al.[16] in a stratified GWAS combining $FI_{adjBMI}$, TG, and HDL cholesterol. These findings confirm that our approach robustly identifies established IR loci, while also nominating 70 novel signals.

**Validation of novel IR loci**

To assess the validity of the 70 novel loci, we examined colocalizations using hypRcoloc (v1.0)[23]. 57 out of 70 (81.4%) of novel variants colocalized with sub-genome-wide TG/HDL associations ($0.05 > P > 5x10^{-8}$, posterior probability range 0.31-1.00; **Supplementary Table 2**). Furthermore, 36 of the 70 novel IR associations (51.4%) overlapped with T2D loci identified by Suzuki et al.[24] (**Supplementary Table 2**), consistent with the role of IR in T2D pathogenesis.

#### Evaluating the pathogenicity of common missense variants

Fine-mapping of IR loci using CARMA[25] identified two missense variants with posterior inclusion probability > 0.8: rs4760 in *PLAUR* (Leu317Pro, PIP=0.83) and rs1133400 in *INPP5A* (Lys45Arg, PIP=0.90) (**Supplementary Table 16**). To assess their potential functional impact, we extracted pathogenicity annotations from the Open Targets Platform[26]. For *PLAUR* Leu317Pro, scores included AlphaMissense[27] = 0.281 (likely benign), GERP [28] = 2.47 (likely conserved), and SIFT[29] = 0 (deleterious). This mixture of bening and deleterious predictions is characteristic of common missense variants, which typically exhibit limited pathogenicity due to purifying selection. For *INPP5A*'s Lys45Arg, predictions were more uniformly benign: AlphaMissense = 0.066 (likely benign), GERP = 0.33 (likely not conserved), and SIFT = 0.28 (tolerated, low confidence).

Structural modeling was available for *INPP5A* Lys45Arg. FoldX's Gibbs energy (ΔΔG)[30,31] predicted a mild destabilizing effect (ΔΔG = 1.28 Kcal/mol). This finding was supported by four additional algorithms: mCSM (ΔΔG = -0.58 kcal/mol)[32], SDM (ΔΔG = -0.08 kcal/mol)[33], DUET (ΔΔG =-0.33 kcal/mol)[34], and DynaMut2 (ΔΔG = -0.90 kcal/mol)[35]. Notably, FoldX computes ΔΔG as $AG_{mutant} – AG_{wild\text{-}type}$ , whereas other methods use the inverse convention. For consistency, in the main text all ΔΔG values are reported following FoldX definition.

**Exploring *LAMB1* expression phenotypic consequences in adipocyte biology:**

*LAMB1* knockdown in SGBS cells[36] increased the number of cells undergoing differentiation, enhanced glucose uptake in preadipocytes (day 0), and increased lipid accumulation in mature adipocytes (day 14) (**Fig. 4C-F**). To expand on these findings, we investigated *LAMB1* expression during adipogenesis in adipose-derived mesenchymal stem cells (AMSCs) from 28 donors[37] (days 0 ,3 ,8 and 14) in basal conditions and under exposure to free fatty acids (FFA). AMSC were profiled by bulkRNAseq and LipocyteProfiler as described in Laber et al[37].

As shown in **Supplementary Fig. 8 and 9,** *LAMB1* expression decreases after day 3. Under basal conditions at day 8, *LAMB1* expression correlated with reduced lipid accumulation and size of large lipid droplets (droplet size as defined in Laber et al[37]). These features positively track adipogenesis (**Supplementary Fig. 8**). These results are consistent with the increased adipogenesis and lipid storage following *LAMB1* knockdown in SGBS cells (**Fig. 4**).

Under FFA exposure, *LAMB1* expression was downregulated in both, immature (day 3) and mature adipocytes (day 14). Given that FFA exposure suppresses adipogenesis[38-40], we hypothesize that *LAMB1* is negatively associated with adipogenic progression, yet requires stage-specific dynamics to support proper differentiation (**Supplementary Fig. 9**).

To examine how *LAMB1* relates to imaging phenotypes under insulin-resistant conditions, we performed linear mixed-effects regression between *LAMB1* expression and LipocyteProfiler[37] features in mature adipocytes (day 14) under FFA exposure adjusting for age, sex BMI, T2D status and batch. *LAMB1* expression (FDR<0.05) was significantly associated with:

- Increased spindle-like morphology

        - Cells_AreaShape_Orientation
- Decreased large lipid droplet-related phenotypes
    - Cells_Mean_LargeBODIPYObjects_Correlation_Overlap_DNA_BODIPY,
    - Cells_Mean_LargeBODIPYObjects_Granularity_2_BODIPY
- Reduced cytoskeletal organization measured with actin, Golgi body and plasma membrane-related (AGP) signal:
    - Cells_Texture_Correlation_AGP_5_03
    - Cytoplasm_Texture_Correlation_AGP_20_00
    - Cells_Texture_Correlation_AGP_5_01
    - Cytoplasm_Texture_Correlation_AGP_5_03
    - Cells_Texture_Correlation_AGP_10_00
    - Cytoplasm_Texture_Correlation_AGP_10_00
    - Cytoplasm_RadialDistribution_RadialCV_DNA_3of4

Taken together, these correlations indicate that higher *LAMB1* expression is associated with impaired lipid accumulation, disrupted cytoskeletal organization, size reduction of large lipid droplets and loss of the characteristic rounded morphology of mature adipocytes[37]. These correlations are consistent with a negative relationship between LAMB1 and adipogenesis.

Next, we computed gene-set enrichment analyses with all genes that co-vary with *LAMB1*-associated phenotypes to assess whether they are enriched for insulin-related biology. 8 out of 10 phenotypes were enriched with at least one gene-set with FDR < 0.05. 4 phenotypes were enriched for lipid metabolism-related pathways:
*Cells_Texture_Correlation_AGP_5_03,Cytoplasm_Texture_Correlation_AGP_20_00,Cells_Texture_Correlation_AGP_10_00 and Cytoplasm_Texture_Correlation_AGP_10_00.*

Of particular interest, genes co-varying with *Cytoplasm_Texture_Correlation_AGP_10_00* are enriched for impaired insulin signaling in adipocytes in both normoglycemic and diabetic conditions (**Supplementary Table 15**). WikiPathways[41] for *Cytoplasm_Texture_Correlation_AGP_10_00*, highlighted focal adhesion pathways and integration of hormone signaling (epithelial growth factor, prolactin, thyroid-stimulating hormone, insulin) (**Supplementary Fig. 10**).

In summary, *LAMB1* expression is negatively associated with adipogenesis and links to pathways regulating insulin signaling, potentially via changes in AGP-related cytoskeletal organization. Moreover, the downregulation of *LAMB1* under FFA exposure suggests that appropriate *LAMB1* expression levels may be required for normal adipogenic progression.

### Supplementary Figures

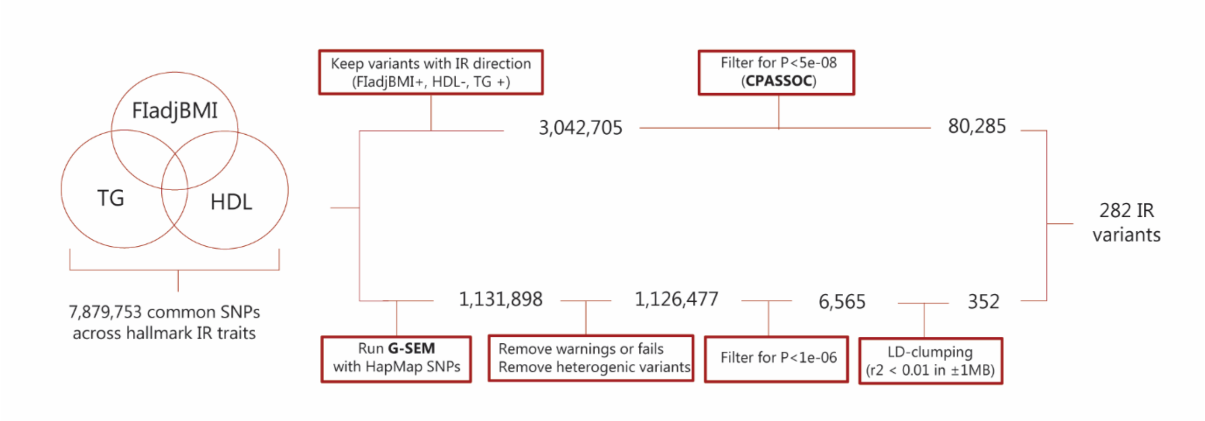


**Supplementary Fig. 1**: Study design for multi-trait identification of IR loci using CPASSOC Shet test[1] and G-SEM common factor GWAS[2]. **CPASSOC**: cross-phenotype association; **$FI_{adjBMI}$**: fasting insulin adjusted for body mass index; **G-SEM**: genomic structural equation modelling; **HDL**: high density lipoprotein cholesterol; **IR**: insulin resistance; **LD**: linkage disequilibrium; **P**: P-value; **SNP**: single nucleotide polymorphism. **TG**: triglycerides.

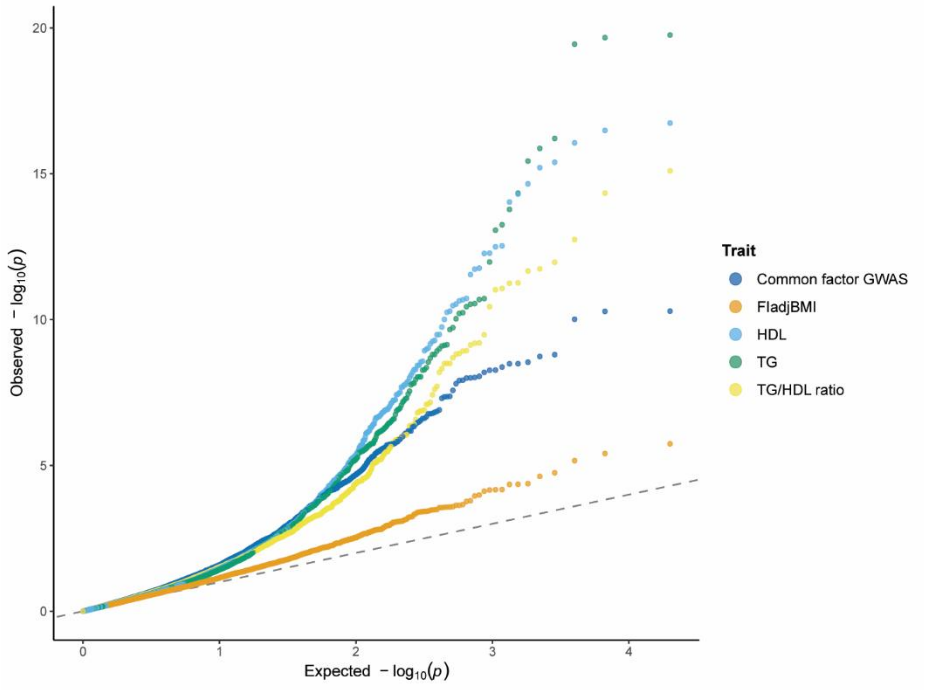


**Supplementary Fig. 2**: QQ-plots for the common factor GWAS and individual insulin resistance traits (TG/HDL ratio[4], and $FI_{adjBMI}$[6], HDL cholesterol[7], and TG[7]). To generate tractable figures, 10,000 associations were randomly sampled for each trait following the approach of Willer et al. [42].

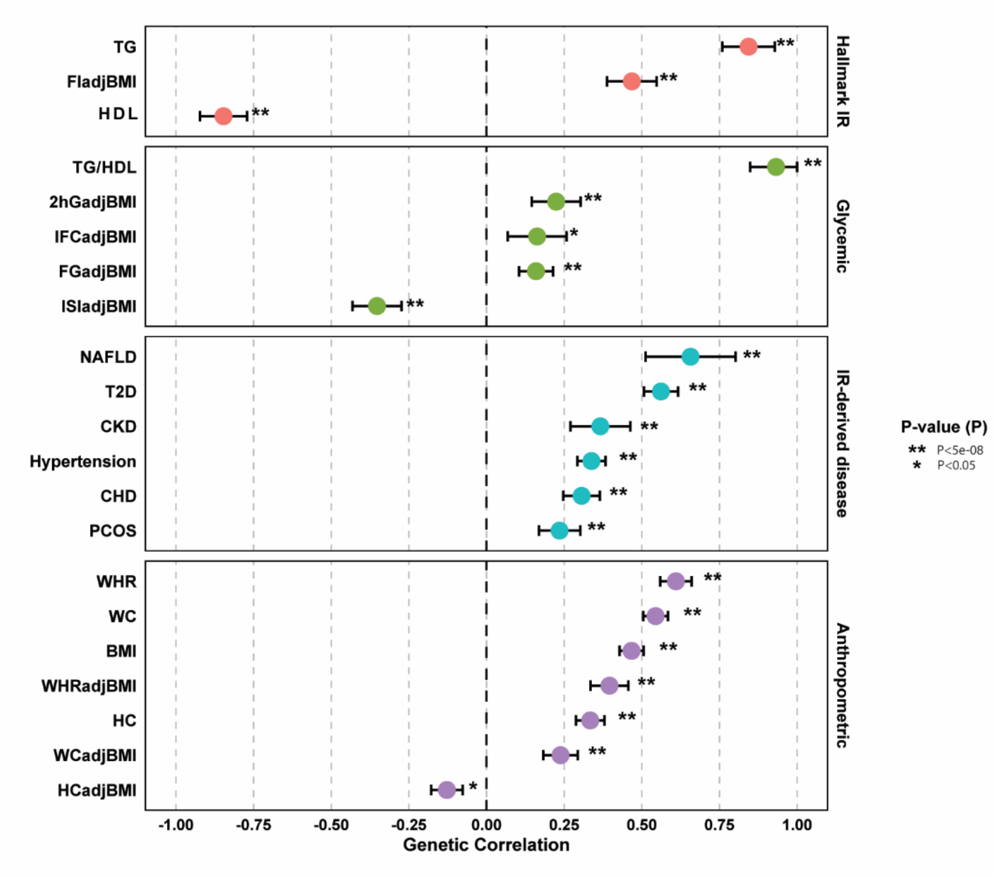


**Supplementary Fig. 3**: Genetic correlations of the common factor GWAS with IR-related traits. Correlations marked with one * indicate nominal significance (P<0.05), and those marked with ** indicate genome-wide significance ($P<5x10^{-8}$). **TG**: triglycerides; **$FI_{adjBMI}$**: fasting insulin adjusted for body mass index; **HDL**: high density lipoprotein cholesterol; **TG/HDL**: triglyceride-to-HDL cholesterol ratio; **$2hG_{adjBMI}$**: two-hour glucose adjusted for body mass index; **$IFC_{adjBMI}$**: insulin-fold change adjusted for body mass index; **$FG_{adjBMI}$**: fasting glucose adjusted for body mass index; **$ISI_{adjBMI}$**: insulin sensitivity index adjusted for body mass index; **NAFLD**: non-alcoholic fatty liver disease; **T2D**: type 2 diabetes; **CKD**: chronic kidney disease; **CHD**: coronary heart disease; **PCOS**: polycystic ovarian syndrome; **WHR**: waist-hip ratio; **WC**: waist circumference; **BMI**: body mass index; **$WHR_{adjBMI}$**: waist-hip ratio adjusted for body mass index; **HC**: hip circumference; **$WC_{adjBMI}$**: waist circumference adjusted for body mass index and **$HC_{adjBMI}$**: hip circumference adjusted for body mass index.

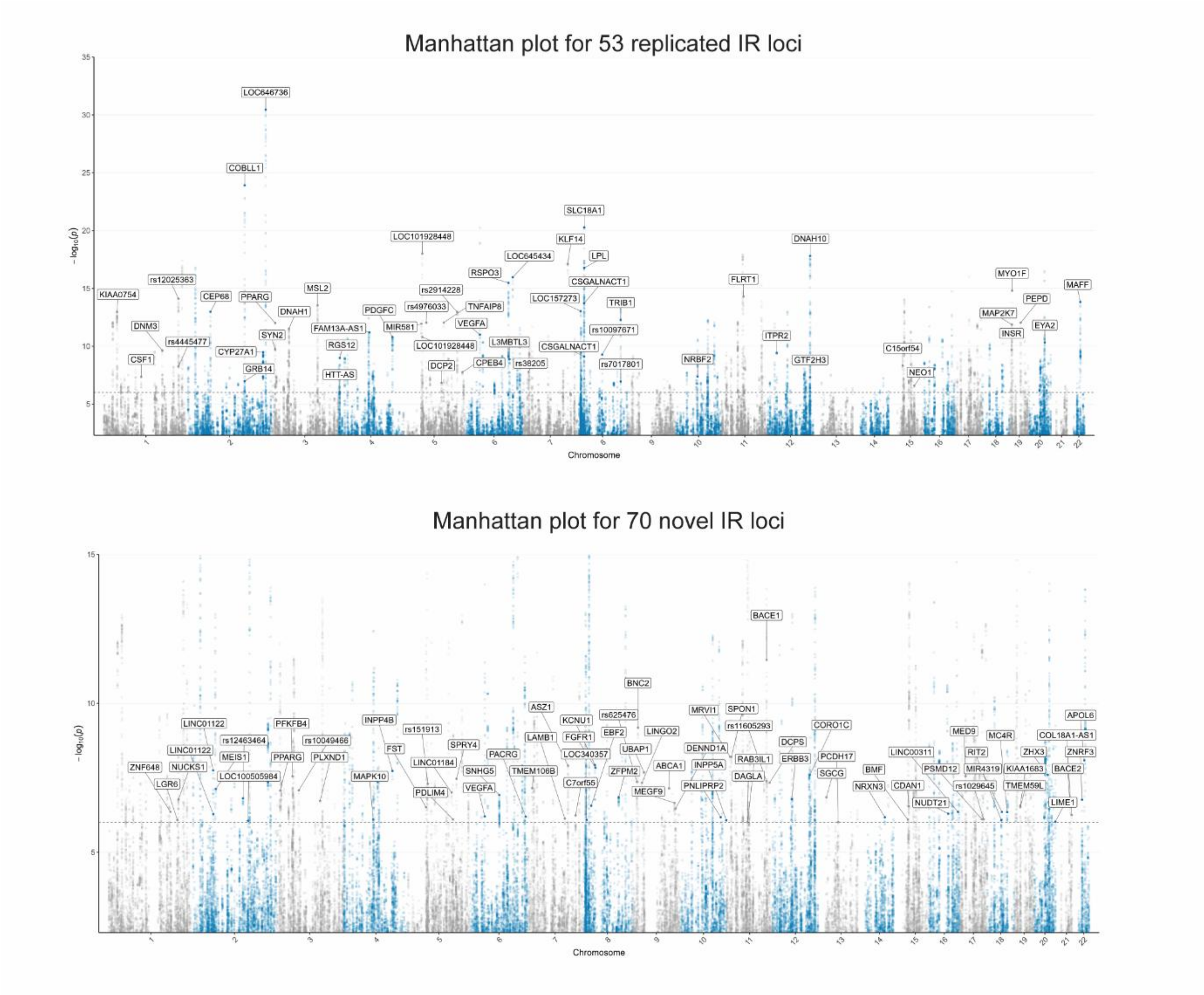


**Supplementary Fig. 4:** Manhattan plots of IR common factor GWAS, highlighting 53 IR signals that replicate previous reported loci from Lotta et al.[16] and 70 novel IR signals identified in this study.

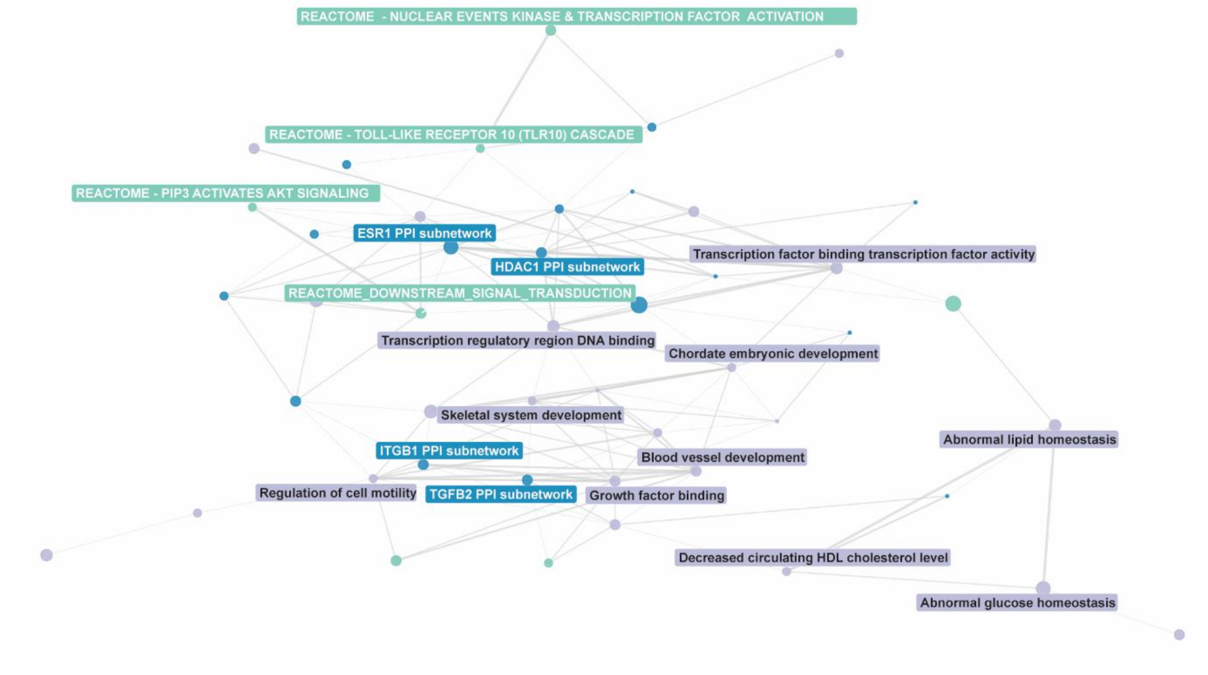


**Supplementary Fig. 5:** Network visualization of significantly enriched gene-sets derived from 282 IR loci using DEPICT[43]. For clarity, only gene sets with Pearson correlation > 0.5 are shown, highlighting clusters of functionally related pathways. **PPI**: protein-protein interactions.

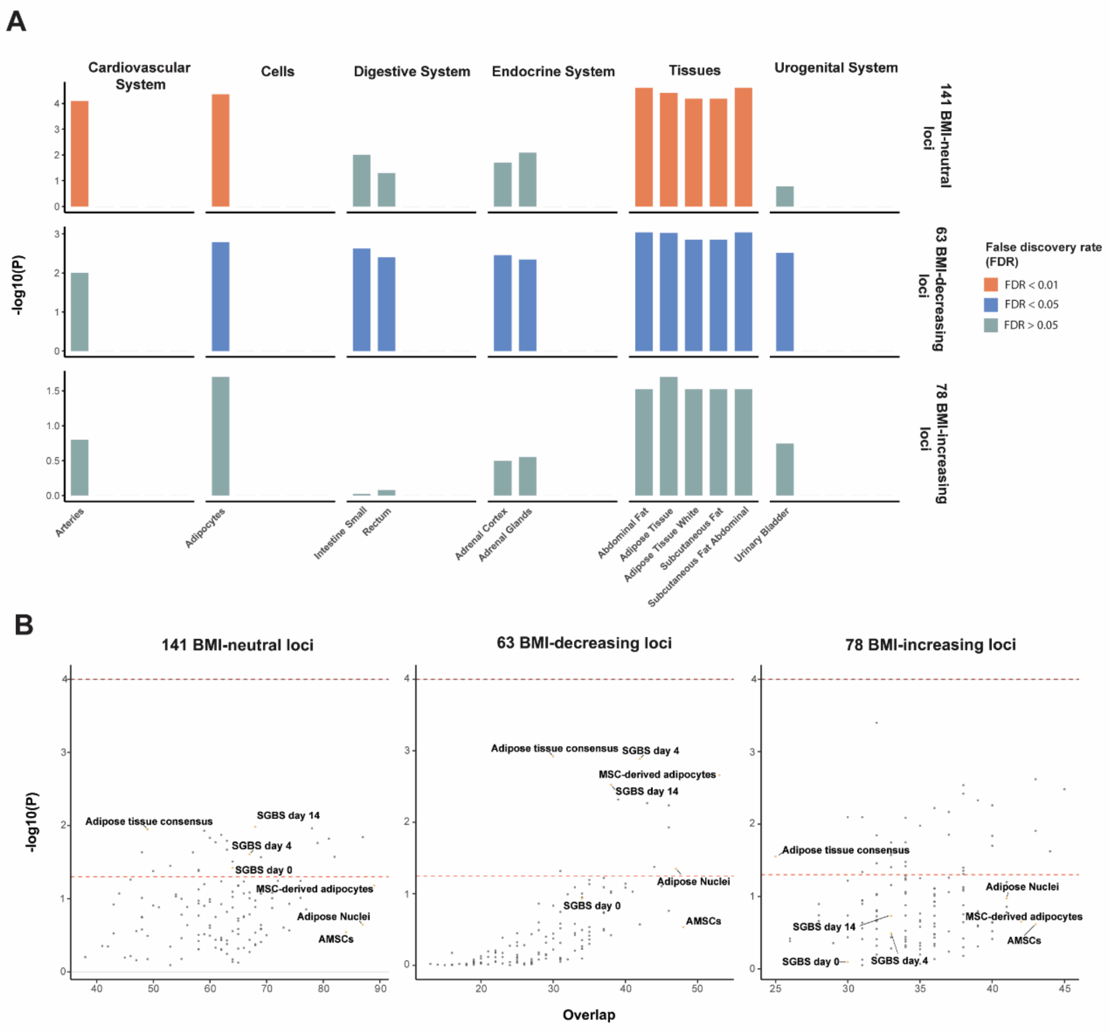


**Supplementary Fig. 6:** Functional enrichment of IR loci by DEPICT[43] and GoShifter[44]. Enrichment analyses for 141 BMI-neutral, 63 BMI-decreasing, and 78 BMI-increasing IR loci. **A.** DEPICT tissue-enrichment results highlighting the top five tissues or cell-types among systems with FDR <0.05 in at least one cluster. **B.** GoShifter enrichment results using (i) enhancer and promoter annotations from the 15-state chromatin model across 127 ROADMAP epigenomes[45], and (ii) ATAC-seq chromatin accessibility peaks from bulk adipose tissue and SGBS cells at days 0, 4 and 14[46]. **AMSCs**: adipose-derived mesenchymal stem cells; **MSCs**: mesenchymal stem cells; **SGBS**: Simpson-Golabi-Behmel Syndrome:

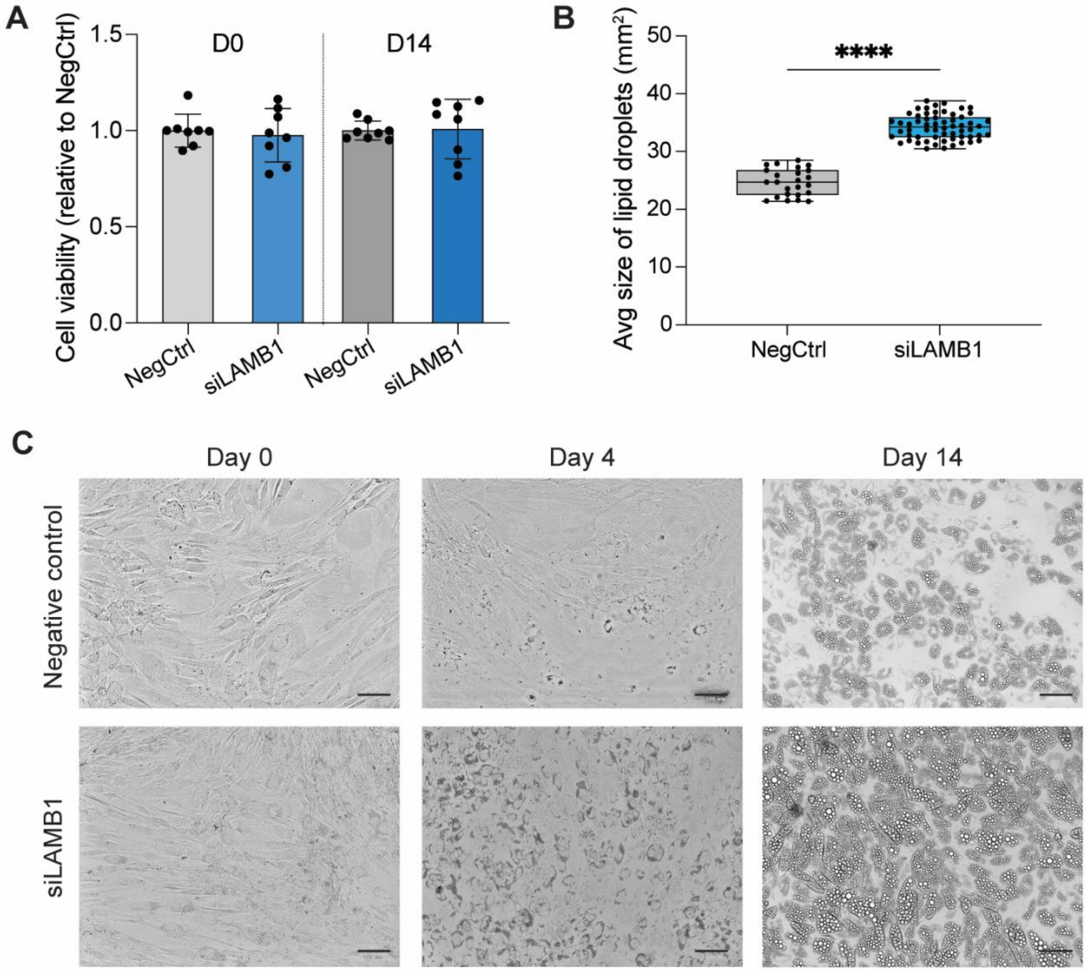


**Supplementary Fig. 7: A.** Cell viability of SBGS cells[36] after siLAMB1 knockdown, normalised to siRNA negative control (NegCtrl) at Day 0 and Day 14 of differentiation. Data are presented as mean ± SD. **B.** Average lipid droplet size measured at Day 14 of differentiation. Data are presented as mean ± SD. **C.** Representative brightfield images of SGBS cells differentiated following siLAMB1 knockdown or negative control treatment at preadipocyte (Day 0), immature adipocyte (Day 4) and mature adipocyte (Day 14) stages. Scale bar = 100 µm.

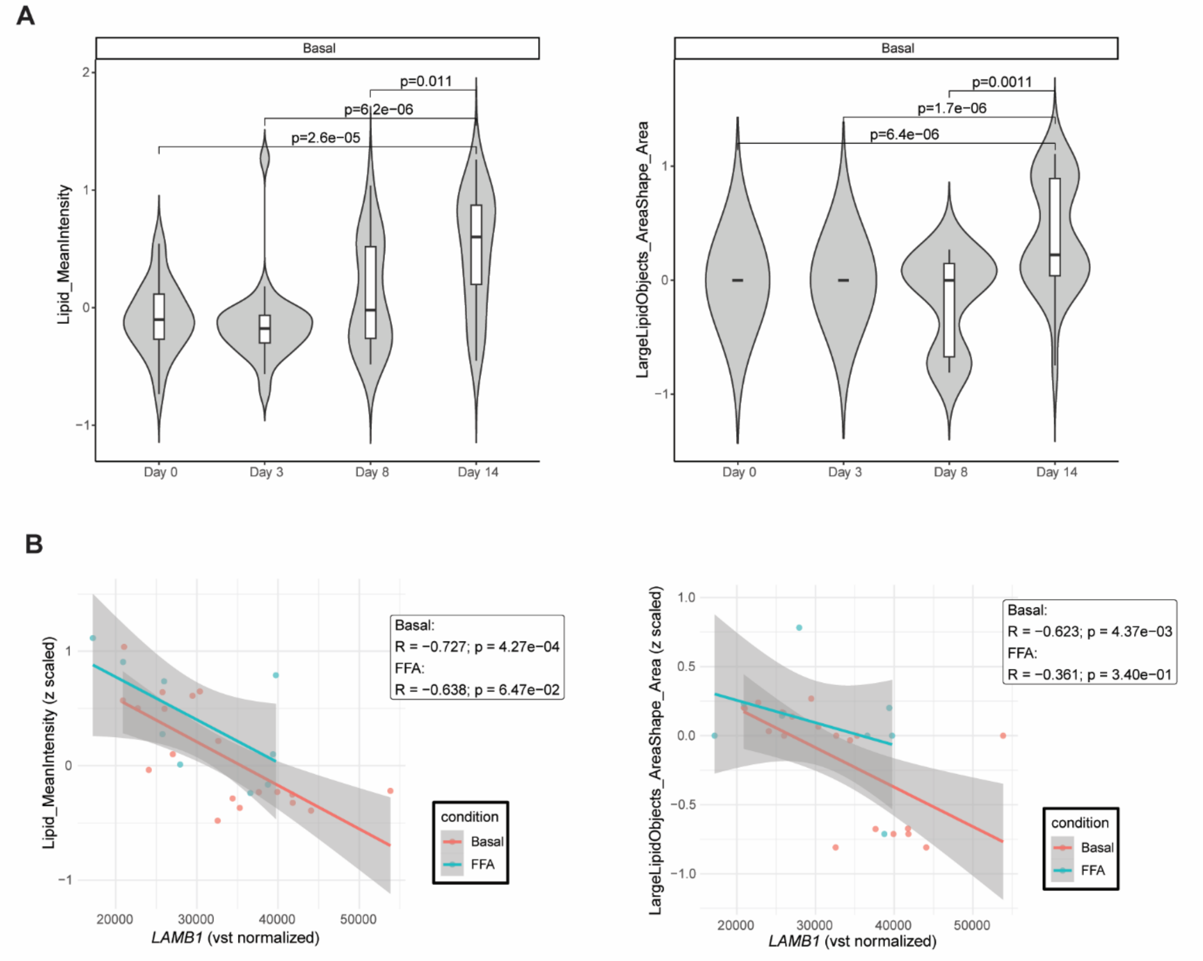


**Supplementary Fig. 8:** Correlation of *LAMB1* expression with phenotypic markers for adipocyte maturity. **A.** Changes in mean intensity of lipid signal (*Lipid_MeanIntensity*) and size of large lipid droplets (*LargeLipidObjects_AreaShape_Area*) in AMSCs[37] across adipogenesis (days 0, 3, 8 and 14). Both measurements progressively increase throughout adipogenesis, with mature adipocytes (day 14) presenting significantly increased signal compared to earlier timepoints (P<0.05) in basal conditions. **B.** *LAMB1* expression is negatively correlated with both mean intensity of lipid signal and size of large lipid droplets. LipocyteProfiler[37] phenotypes are z-scaled and *LAMB1* expression is variance-stabilized (vst) normalized to facilitate comparability across samples. **AMSCs**: adipose mesenchymal stem cells; **FFA**: free fatty acid; **Z**: z-score; **P**: P-value; **vst**: variance-stabilizing transformation.

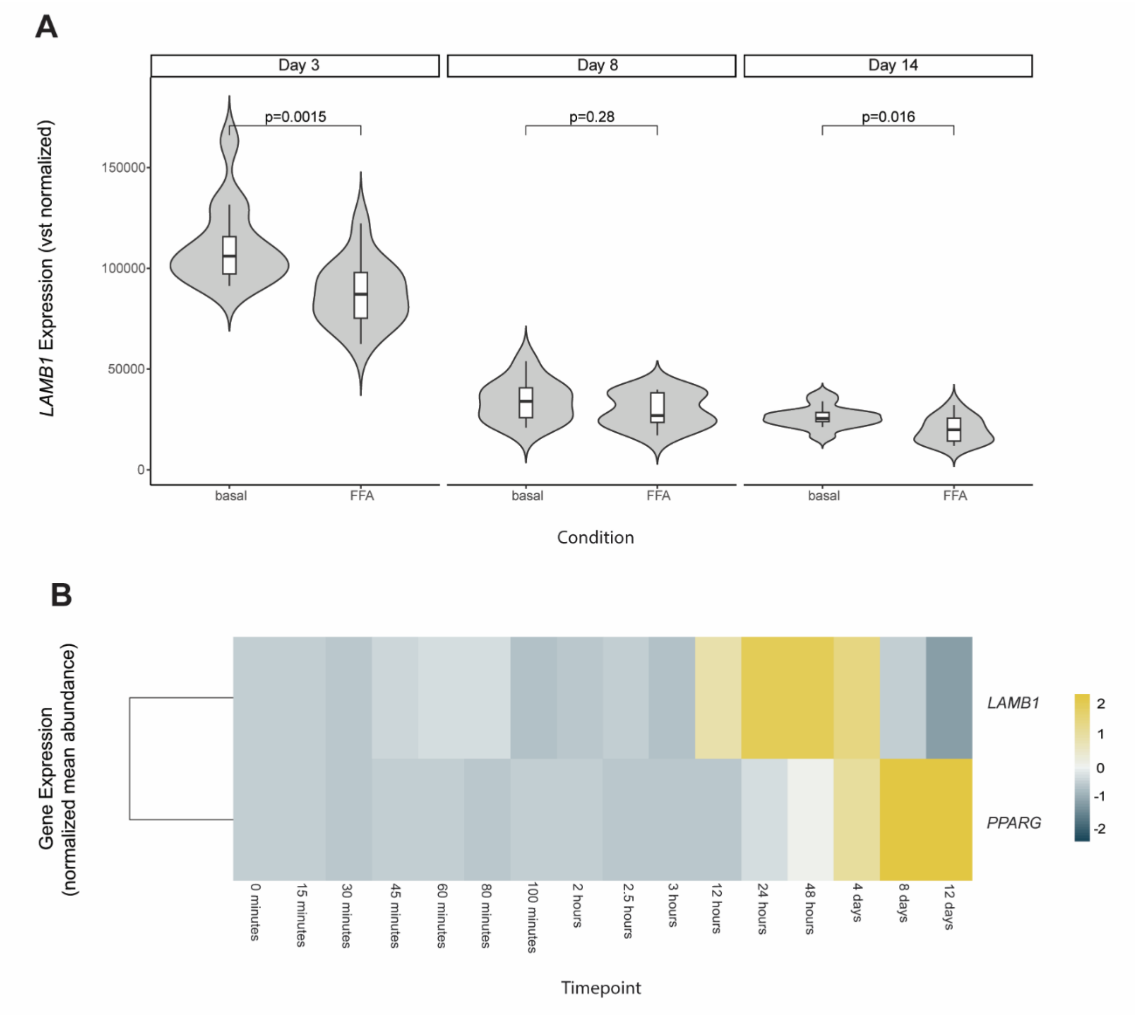


**Supplementary Fig. 9:** *LAMB1* expression dynamics in AMSCs[37] across adipogenesis under basal and FFA-exposed conditions. **A.** Under FFA exposure *LAMB1* expression is significantly decreased at days 3 and 14 compared to basal conditions (P<0.05). **B.** Reference trajectory of *LAMB1* expression across adipogenesis in AMSCs from adiposetissue.org[47]. For comparison, *PPARG* expression is also shown. *LAMB1* expression starts at day 0 and peaks in immature adipocytes (day 4) and decreases in mature adipocytes (days 8 and 12). **AMSCs**: adipose mesenchymal stem cells; **FFA**: free fatty acid; **P**: P-value; **vst**: variance-stabilizing transformation.

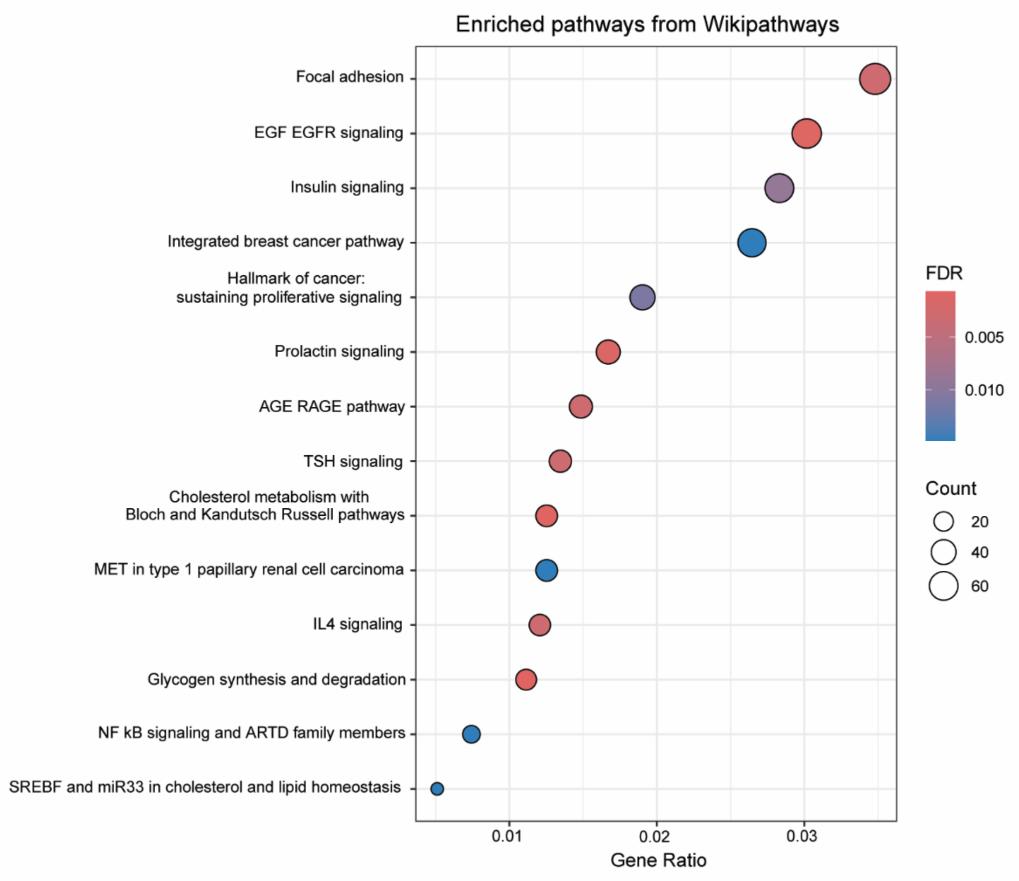


**Supplementary Fig. 10:** Pathway enrichment results for genes that co-vary with LipocyteProfiler-derived, *LAMB1*-associated phenotype *Cytoplasm_Texture_Correlation_AGP_10_00.* All Wikipathways[41] terms that are significantly enriched at FDR<0.05 are shown. Pathways are ordered by descending gene ratio, reflecting the number of phenotype-associated genes annotated to each pathway (Count). **AGE**: advanced glycation end-products; **ARTD**: ADP-Ribosyltransferase D; **EGF**: epidermal growth factor; **EGFR**: EGF receptor; **IL4**: interleukin 4; **MET**: mesenchymal-epithelial transition; **miR33**: microRNA-33; **NF-kb**: Nuclear Factor kappa-light-chain-enhancer of activated B cells; **RAGE:** Receptor for AGE; **SREBF**: Sterol Regulatory Element Binding Factor**; TSH**: thyroid stimulating hormone.

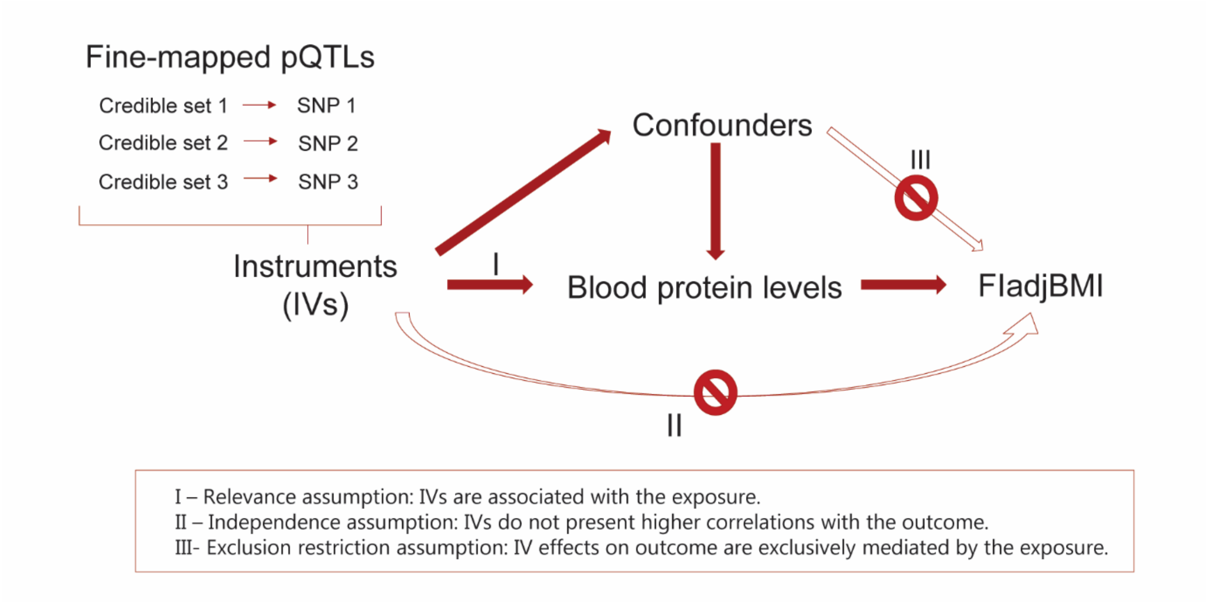


**Supplementary Fig. 11:** Schematic overview of the Mendelian randomization framework applied in this study, with pQTLs serving as instrumental variables (IVs). The figure also summarizes the core MR assumptions required for causal inference. pQTL: protein quantitative trait locus; $FI_{adjBMI}$: fasting insulin adjusted for body mass index.

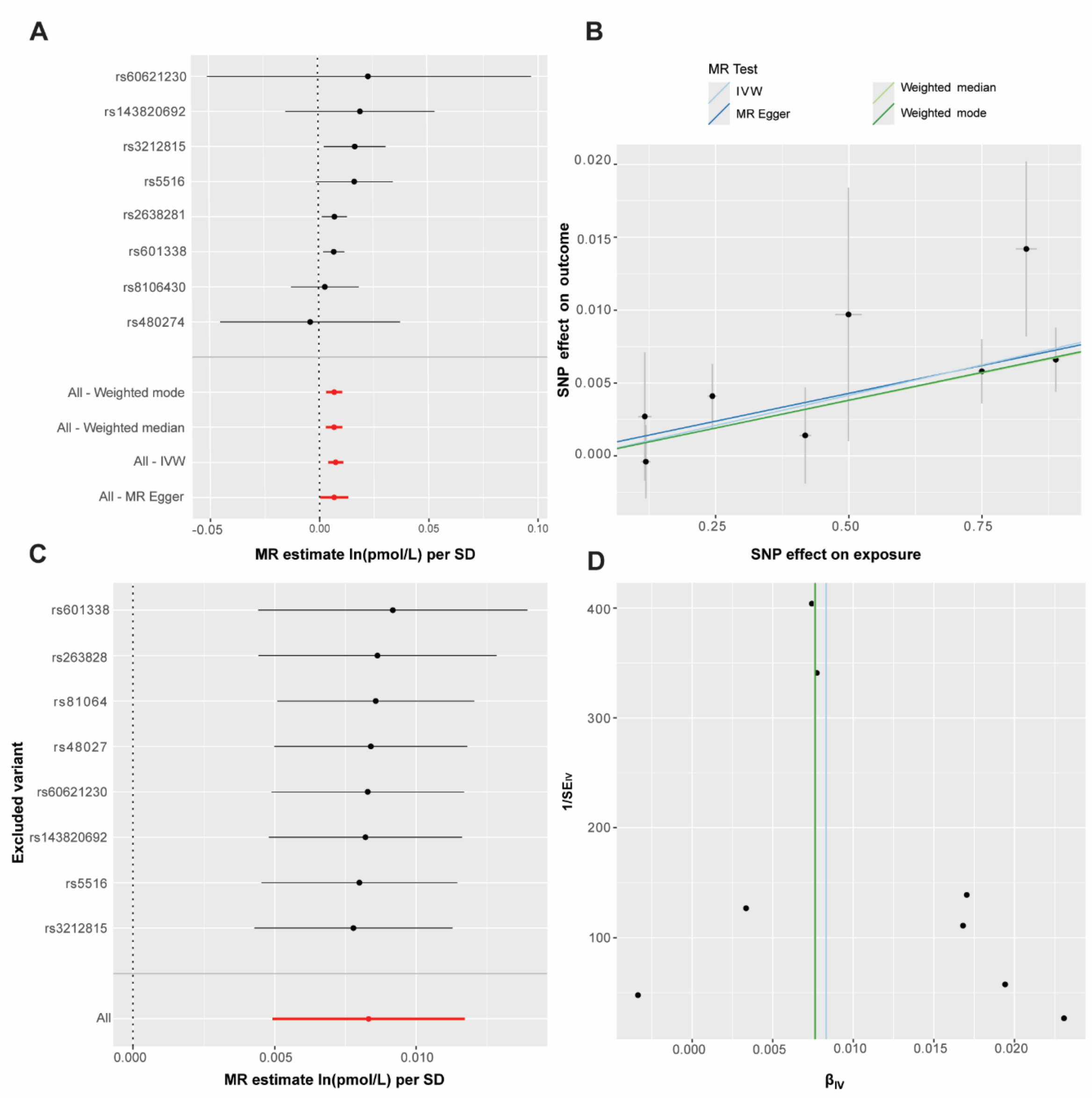


**Supplementary Fig. 12:** Diagnostic plots for Mendelian Randomization analyses testing the causal relationship between KLK1 protein levels and $FI_{adjBMI}$. **A).** Forest plot of individual instrument estimates. **B.** Scatter plot of genetic associations with exposure and outcome. **C.** Leave-one out plot assessing robustness to single-variant exclusion. **D.** Funnel plot evaluating symmetry and potential directional pleiotropy. **IV**: instrument variant; **IVW**: inverse variance weighted; **MR**: mendelian randomization; **pmol/L**: picomoles per litre; **SNP**: single nucleotide polymorphism.
